# Safety and immunogenicity of a broad-spectrum mosaic vaccine as a booster dose against SARS-CoV-2 Omicron and other circulating variants

**DOI:** 10.1101/2022.09.05.22279589

**Authors:** Nawal Al Kaabi, Yun Kai Yang, Yu Liang, Ke Xu, Xue Feng Zhang, Yun Kang, Yu Qin Jin, Jun Wei Hou, Jing Zhang, Tian Yang, Salah Hussein, Mohamed Saif ElDein, Ze Hua Lei, Hao Zhang, Shuai Shao, Zhao Ming Liu, Ning Liu, Xiang Zheng, Ji Guo Su, Sen Sen Yang, Xiangfeng Cong, Yao Tan, Wenwen Lei, Xue Jun Gao, Zhiwei Jiang, Hui Wang, Meng Li, Hanadi Mekki Mekki, Walid Zaher, Sally Mahmoud, Xue Zhang, Chang Qu, Dan Ying Liu, Jing Zhang, Mengjie Yang, Islam Eltantawy, Peng Xiao, Fu Jie Shen, Jin Juan Wu, Zi Bo Han, Li Fang Du, Fang Tang, Shi Chen, Zhi Jing Ma, Fan Zheng, Ya Nan Hou, Xin Yu Li, Xin Li, Zhao Nian Wang, Jin Liang Yin, Xiao Yan Mao, Jin Zhang, Liang Qu, Yun Tao Zhang, Xiao Ming Yang, Guizhen Wu, Qi Ming Li

**Affiliations:** Sheikh Khalifa Medical City, SEHA, Abu Dhabi, United Arab Emirates; College of Medicine and Health Sciences, Khalifa University, Abu Dhabi, United Arab Emirates; China National Biotec Group Company Limited, Beijing, China; The Sixth Laboratory, National Vaccine and Serum Institute (NVSI), Beijing, China; National Engineering Center for New Vaccine Research, Beijing, China; National Institute for Viral Disease Control and Prevention, Chinese Center for Disease Control and Prevention (China CDC), Beijing, China; Clinical Medicine Office, National Vaccine and Serum Institute (NVSI), Beijing, China; Lanzhou Institute of Biological Products Company Limited, Lanzhou, China; Beijing Key Tech Statistical Consulting Co., Ltd, Beijing, China; Beijing Institute of Biological Products Company Limited, Beijing, China; Union 71, United Arab Emirates; G42 Healthcare, United Arab Emirates

## Abstract

**BACKGROUND:** The rising breakthrough infections caused by severe acute respiratory syndrome coronavirus 2 (SARS-CoV-2) variants, especially Omicron and its sub-lineages, have raised an urgent need to develop broad-spectrum vaccines against [coronavirus disease 2019 (COVID-19)](https://www.bing.com/ck/a?!&&p=7a72ff758ab318d6JmltdHM9MTY2MTQxMTI5NSZpZ3VpZD03Y2QxMWU0MS1jNGNlLTRjNzAtYjA3Yi0xMTVjZmM1N2RlODQmaW5zaWQ9NTE2Ng&ptn=3&hsh=3&fclid=b014239e-2444-11ed-959d-82aad9d02527&u=a1aHR0cHM6Ly93d3cuY2RjLmdvdi9jb3JvbmF2aXJ1cy8yMDE5LW5jb3YvaW5kZXguaHRtbA&ntb=1). We have developed a mosaic-type recombinant vaccine candidate, named NVSI-06-09, having immune potentials against a broad range of SARS-CoV-2 variants.

**METHODS:** An ongoing randomized, double-blind, controlled phase 2 trial was conducted to evaluate the safety and immunogenicity of NVSI-06-09 as a booster dose in subjects aged 18 years and older from the United Arab Emirates (UAE), who had completed two or three doses of BBIBP-CorV vaccinations at least 6 months prior to the enrollment. The participants were randomly assigned with 1:1 to receive a booster dose of NVSI-06-09 or BBIBP-CorV. The primary outcomes were immunogenicity and safety against SARS-CoV-2 Omicron variant, and the exploratory outcome was cross-immunogenicity against other circulating strains.

**RESULTS:** A total of 516 participants received booster vaccination. Interim results showed a similar safety profile between NVSI-06-09 and BBIBP-CorV booster groups, with low incidence of adverse reactions of grade 1 or 2. For immunogenicity, by day 14 after the booster vaccination, the fold rises in neutralizing antibody geometric mean titers (GMTs) from baseline level elicited by NVSI-06-09 were remarkably higher than those by BBIBP-CorV against the prototype strain (19.67 vs 4.47-fold), Omicron BA.1.1 (42.35 vs 3.78-fold), BA.2 (25.09 vs 2.91-fold), BA.4 (22.42 vs 2.69-fold), and BA.5 variants (27.06 vs 4.73-fold). Similarly, the neutralizing GMTs boosted by NVSI-06-09 against Beta and Delta variants were also 6.60-fold and 7.17-fold higher than those boosted by BBIBP-CorV.

**CONCLUSIONS:** A booster dose of NVSI-06-09 was well-tolerated and elicited broad-spectrum neutralizing responses against SARS-CoV-2 prototype strain and immune-evasive variants, including Omicron and its sub-lineages. The immunogenicity of NVSI-06-09 as a booster vaccine was superior to that of BBIBP-CorV. (Funded by LIBP and BIBP of Sinopharm; ClinicalTrials.gov number, NCT05293548).

## INTRODUCTION

Severe acute respiratory syndrome coronavirus 2 (SARS-CoV-2) continuously evolves to acquire mutations that may change its infectivity and antigenicity. Since the beginning of SARS-CoV-2 pandemic, several variants with increased transmissibility or immune escape capability have emerged, such as Beta, Gamma, Delta and Omicron.^1-3^ These variants have caused successive waves of SARS-CoV-2 infections and increasing numbers of breakthrough cases.^4,5^ Especially, the recently emerged Omicron is the most antigenically divergent variant, known to date, from the ancestral strain, which consists of several distinct sublineages including BA.1, BA.2, BA.3, BA.4 and BA.5.^6-8^ Owing to their much higher transmissibility and greater ability to evade prior immunity, Omicron and its sublineages spread rapidly around the world even in the background of high vaccination and previous infection rates in populations.^6-9^ The rapid emergence of new immune-evasive SARS-CoV-2 variants has seriously threatened the efficacy of vaccines currently used, and highlights the urgent need of next-generation vaccines with broad-spectrum protections against divergent SARS-CoV-2 variants.

Guided by structural and computational analyses, we have designed a trimeric RBD vaccine, named NVSI-06-07, in which three homologous RBDs derived from the prototype SARS-CoV-2 strain were connected end-to-end and co-assembled into a trimeric structure. Clinical studies demonstrated that a booster dose of NVSI-06-07 following primary vaccination of the inactivated vaccine significantly improves the neutralizing antibody (nAb) responses against SARS-CoV-2.^10,11^ NVSI-06-07 has been approved by the United Arab Emirates (UAE) for emergency use. The trimeric RBD that accommodates three RBDs in a single immunogen facilitates us to design a mosaic-type vaccine (NVSI-06-09), which integrates the key mutations from Omicron and other circulating variants into one molecule. Pre-clinical studies in animals showed that NVSI-06-09, either used alone or as a booster dose, induced potent and broad immune responses against divergent SARS-CoV-2 variants including Omicron, which may potentially serve as a broad-spectrum vaccine candidate.^12^

Here we reported the interim analysis results of a randomized, double-blind, controlled phase 2 trial conducted in UAE to evaluate the safety and immunogenicity of NVSI-06-09 as a booster dose in the inactivated vaccine BBIBP-CorV recipients, using the homologous boost with one additional dose of BBIBP-CorV as the control arm. Considering the dominant prevalence of Omicron worldwide currently, immune responses against multiple Omicron sub-lineages were evaluated. Furthermore, as an exploratory study, the cross-neutralizing antibody responses against other variants of concern (VOCs) were also detected to assess the broad-spectrum immune responses elicited by NVSI-06-09.

## METHODS

### TRIAL DESIGN

We conducted a randomized, double-blind, controlled phase 2 trial to assess the safety and immunogenicity of the mosaic-type trimeric RBD vaccine, named NVSI-06-09, as a heterologous booster shot following primary vaccination of an inactivated vaccine BBIBP-CorV. The homologous boost of another dose of BBIBP-CorV was used as control in the study. The trial was reviewed and approved by the Abu Dhabi Health Research and Technology Ethics Committee of UAE, and performed in accordance with the principles of the International Conference on Harmonization-Good Clinical Practice (ICH-GCP) and the Declaration of Helsinki (with amendments). This study also met the local legal and regulatory requirements of UAE. The sponsors of the study are China National Biotec Group Co., Ltd. (CNBG) of Sinopharm, National Vaccine and Serum Institute (NVSI) of Sinopharm CNBG, Lanzhou Institute of Biological Products Co., Ltd. (LIBP) of Sinopharm CNBG, and Beijing Institute of Biological Products Co., Ltd. (BIBP) of Sinopharm CNBG. The study protocol is available at NEJM.org.

### PARTICIPANTS

Eligible trial participants were healthy men and nonpregnant women, aged 18 years or above, who had previously vaccinated with two or three doses of BBIBP-CorV at least six months prior to enrollment. Written informed consent was obtained from all volunteers. During screening, healthy status of each volunteer was assessed by inquiry and physical examination. Volunteers who had previously received any COVID-19 vaccine other than BBIBP-CorV were excluded. Individuals whose axillary temperature ≥37.3 °C (or forehead temperature ≥37.8 °C) or allergic to any components of the vaccine were also excluded. Other exclusion criteria included history of thrombocytopenia or other coagulation disorders; immunological impairment or immunocompromise; receipt of any blood products or immunoglobulin therapy within three months prior to enrollment; serious chronic disease; receipt of any other inactivated vaccines in the past 14 days, or any live attenuated vaccines post one month; receipt of any investigational drugs within 6 months prior to enrollment; and other vaccination-related contraindications considered by the investigator.

### STUDIED VACCINES

NVSI-06-09 is a mosaic-type trimeric RBD vaccine, recombinantly expressed by Chinese hamster ovary (CHO) cells, which covalently combined three heterologous RBDs into a single molecule to present potentially broad immunological coverage. Within NVSI-06-09, one RBD is derived from Omicron variant, and the other two are artificially designed harboring the key residues from other variants of concern (VOCs) and variants of interests (VOIs).^12^ This vaccine was developed by the NVSI of Sinopharm CNBG, and provided in the liquid form at a single dose of 0.5 ml/vial, containing 20 μg antigen protein, 0.3 mg aluminum hydroxide, 0.39 mg histidine and 4.38 mg sodium chloride for injection. The inactivated vaccine BBIBP-CorV, used as a control vaccine in this study, was developed by the BIBP of Sinopharm CNBG using the prototype SARS-CoV-2 virus (19nCoV-CDC-Tan-HB02 strain). BBIBP-CorV is manufactured by culturing virus in Vero cells and inactivated by β-propionolactone,^13^ and provided in a single dose of 0.5 ml per vial containing 6.5 U antigen. BBIBP-CorV is one of the COVID-19 vaccines authorized by the World Health Organization (WHO) and has been widely used in many countries around the world. Both NVSI-06-09 and BBIBP-CorV are transported and stored at 2 – 8°C.

### RANDOMIZATION AND BLINDING

In this study, the participants were randomly assigned in a ratio of 1:1 to receive either a heterologous booster dose of NVSI-06-09 or a homologous booster dose of BBIBP-CorV. For participant allocation, eligible persons were assigned to two booster groups using a stratified blocked randomization method by SAS 9.4 software, in which stratification was based on different doses (two or three doses) of BBIBP-CorV that they received prior to enrollment and the block size was set to 4. The vaccine randomization list was also generated by SAS 9.4 software using the block randomization method with a block size of 4. Both participant and vaccine randomization lists were generated by an unblinded statistician, and then imported into the Interactive Web Response System (IWRS). After enrollment, each participant was assigned a randomization number from IWRS. At the clinical site, a vaccine number was also obtained from IWRS for vaccination accordingly. If the vaccine vial is damaged, a new vaccine number will be assigned to the participant.

Participants and clinical operation team involved in safety data collection and immunogenicity assessments were blind to treatment allocation during the trial. Vaccine preparation was done by independent personnel to ensure identical appearance between the studied and control vaccines. Individuals involved in randomization and blinding did not participate in other trial operations.

### PROCEDURES

At the clinical site, eligible participants were randomly assigned to receive a booster vaccination with NVSI-06-09 or BBIBP-CorV via intramuscular injection into the lateral deltoid muscle of the upper arm. After vaccination, participants were observed in the observation room for at least 30 minutes for any immediate adverse reactions. Solicited local adverse events at injection site (including pain, pruritus, induration, swelling, rash, and redness) and solicited systemic adverse events (including non-vaccination site muscle pain, headache, fever, fatigue, pruritus at non-vaccination site, arthralgia, constipation, vomiting, nausea, cough, dyspnea, dizziness, diarrhea, dysphagia, anorexia, abnormal skin mucosa, and acute allergic reaction) were recorded within 7 days post-vaccination. Unsolicited adverse events were collected within 30 days after the receipt of vaccination. Serious adverse events and the adverse events of special interest were monitored for 12 months after vaccination. All the reported adverse events were reviewed and verified by investigators, and the grades of adverse events were determined based on the relevant guidance of China National Medical Products Administration (NMPA). Blood samples were taken from participants for immunogenicity assessment on day 0, day 14, day 28, and at 3 months, 6 months, 9 months, and 12 months, respectively.

### STUDY OUTCOMES

The primary outcomes were the immunogenicity and safety of the booster vaccination of NVSI-06-09 against SARS-CoV-2 Omicron variant, in comparison with the boost of BBIBP-CorV, on day 14 after the vaccination. The immunogenicity was evaluated by the geometric mean titers (GMTs) of anti-Omicron neutralizing antibodies (nAbs), and the four-fold rise rate of the neutralizing GMTs from baseline. Anti-Omicron nAb titers were measured by live-virus microneutralization assays. The safety was assessed by occurrence and severity of adverse reactions. NVSI-06-09 was designed as a potential broad-spectrum COVID-19 vaccine. To assess the broad immune responses induced by a booster dose of NVSI-06-09, the cross-neutralizing antibody GMTs against other SARS-CoV-2 strains, including the prototype, Beta and Delta, were also detected as an exploratory study.

### LABORATORY TESTS

NAb titers in the sera of participants were detected by using the live-virus microneutralization assay based on inhibition of cytopathic effects (CPE). A total of six SARS-CoV-2 live viruses, including Omicron BA.1.1 (NPRC 2.192100005), Omicron BA.2 (NPRC 2.192100010), Omicron BA.4 (NPRC 2.192100012), Omicron BA.5 (NPRC 2.192100015), prototype (QD-01), Beta (GD84) and Delta (GD96) strains, were tested. These viruses were provided by the National Institute for Viral Disease Control and Prevention, the Chinese Center for Disease Control and Prevention (China CDC), Beijing, China. The live-virus microneutralization assays were also performed in the Biosafety Level-3 Laboratory (BSL-3) of the National Institute for Viral Disease Control and Prevention, China CDC.

In the CPE-based live-virus microneutralization assay, serum samples were firstly heat-inactivated at 56 °C for 30 min, followed by a series of two-fold dilutions starting from 1:4 for pre-booster serum samples and 1:8 for post-booster samples. The diluted serum was mixed with an equal volume of SARS-CoV-2 solution containing 100 TCID_50_ of live virus in the wells on the 96-well plate. Then the plate was incubated at 37°C with 5% carbon dioxide (CO_2_) for 2 hours. Subsequently, Vero cells, cultured in the medium 199 containing 5% fetal bovine serum (FBS), with a density of (1.0-2.0)×10^5^ per mL were added to the wells, and incubated at 37 °C±1°C with 5% CO_2_ for 5 ∼ 7 days. After incubation, cytopathic changes in Vero cells were examined, and nAb titers were determined as the reciprocal of serum dilution at which 50% of CPE was inhibited. The titer of the serum below the limit of quantification was reported as the half value of the quantification limit. In the assay, both negative and positive reference sera were tested as controls. The reference sera and Vero cells were provided by the National Institute for Food and Drug Control (NIFDC) of China. Cell-control was also set and the titer of the virus was also re-titrated in the assay.

### STATISTICAL ANALYSIS

The sample size of trial participants was determined using the Power Analysis and Sample Size (PASS15.0) software. It was assumed that the anti-Omicron nAb GMT in the experimental group was superior to that in the control group on day 14 after vaccination, with a superiority margin after log10 transformation of 0.0212. The other assumption parameters include standard deviation of the antibody GMT after log10 transformation of 0.55, the type I error (one-sided) probability of 0.025, the expected power of 0.9, and a 20% dropout rate. As such, a total of 398 participants were required. It was further assumed that the nAb level in the experimental group was non-inferior to that in the control group on day 14 post-vaccination. When using the non-inferiority threshold after log10 transformation of −0.17609 and the other parameters remained the same as above, a total of 516 participants were required to achieve 0.9 power to detect non-inferiority. Taken together, the sample size was determined to be 516, with 258 in the experimental group and the other 258 in the control group.

Baseline characteristics were analyzed based on the full analysis set (FAS), which includes all randomized participants whose baseline data were available and valid. Safety analysis was performed on the safety set (SS) that includes all participants who received the booster vaccination. Immunogenicity was analyzed based on the per-protocol set (PPS) including all participants who received the booster vaccination and had valid pre- and post-vaccination immunogenicity data.

For baseline characterization, continuous and categorical characteristics between groups were compared using two-sided Student’s t-test and two-sided Chi-square test, respectively. In the safety analysis, both counts and percentage of adverse reactions were presented, and two-sided Fisher’s exact test was used to determine whether there was a significant difference between the safety of two groups. In immunogenicity analysis, the GMT of nAbs, the fold rise of the neutralizing GMT from baseline and the ratio of GMTs between the experimental and control groups, and the associated 95% confidence interval (CI) were calculated. In addition, the four-fold rise rates of nAb titers from baselines and 95% CIs were calculated by the Clopper–Pearson method. The differences in the four-fold rise rates between the experimental and control groups as well as the associated 95% CIs were evaluated by Cochran–Mantel–Haenszel (CMH) method considering stratification factors. The adjusted nAb GMT and the corresponding *p* value were calculated using covariance analysis with least square method. The nAb GMTs between two groups were compared using the two-sided grouped t-test after log10 transformation of the titers, and the difference in the neutralizing GMT fold rise between two groups was also analyzed using the two-sided grouped t-test. All the above analyses were performed using the SAS (version 9.4) software.

## RESULTS

### PARTICIPANTS AND BASELINE DEMOGRAPHIC CHARACTERISTICS

Between May 25 and 30, 2022, a total of 522 healthy adults aged 18 years and older, who had received vaccinations of the inactivated vaccine BBIBP-CorV at least 6 months prior, were screened and enrolled. After enrollment, six individuals withdrew from the study and 516 participants received the booster dose of vaccination (Fig. 1). Among these 516 participants, 21 were administered with two doses of BBIBP-CorV and 495 with three doses prior to the study. The participants were randomly assigned to receive a booster dose of NVSI-06-09 (N=260) or a booster vaccination of BBIBP-CorV (N=256). All the 516 participants were included in the full analysis set (FAS) for baseline characterization and in the safety set (SS) for safety assessment. A total of 504 participants (N=255 with NVSI-06-09 and N=249 with BBIBP-CorV) who had valid immunogenicity results without protocol deviation were included in the per-protocol set (PPS) for immunogenicity assessment (Fig. 1).

**Figure 1.**
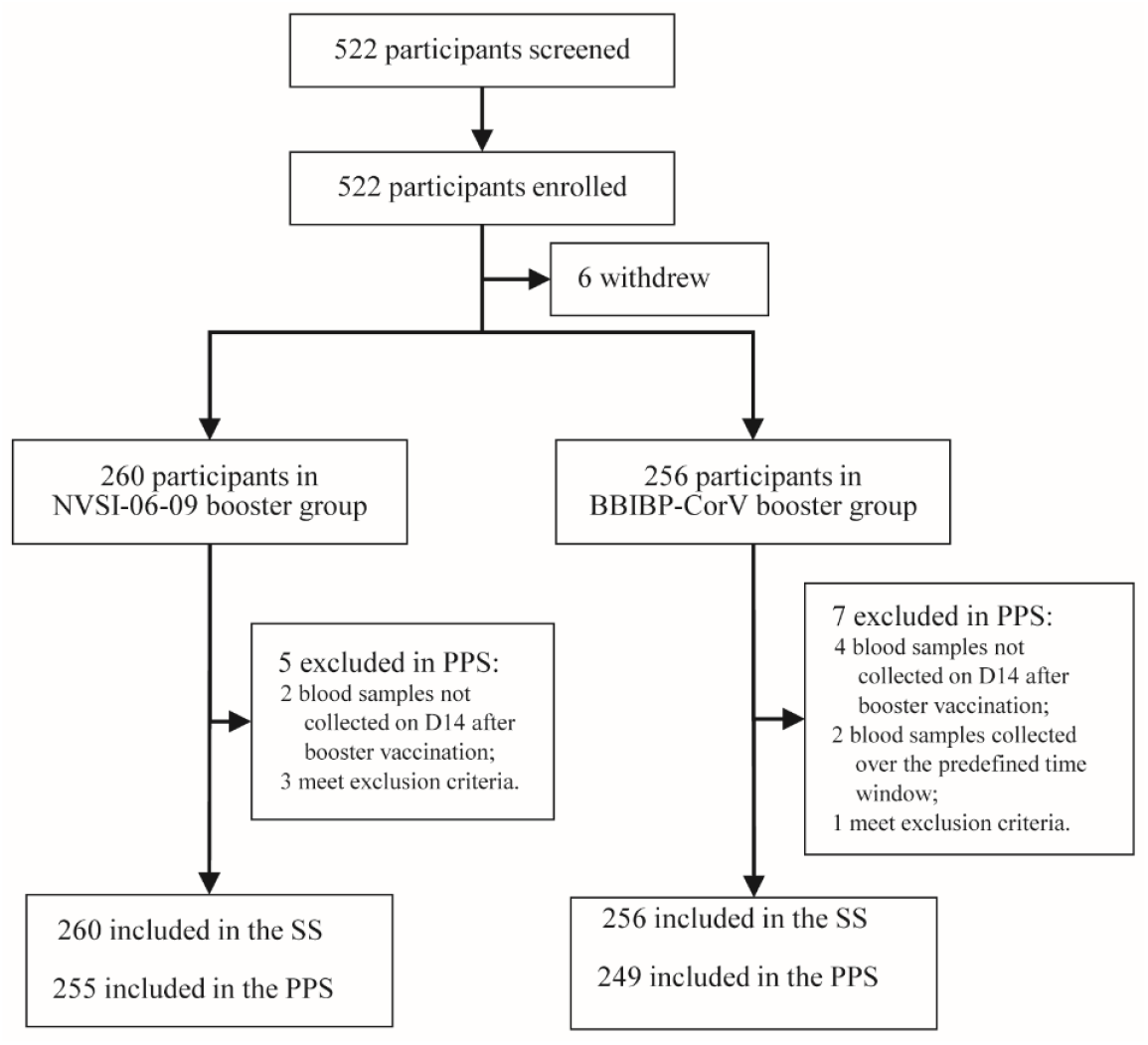
Screening, randomization, and analysis populations. 522 healthy adults were screened and enrolled, of which 6 persons withdrew before vaccination and 516 participated in the study. The participants were randomly assigned to receive a booster injection of either NVSI-06-09 (260 participants) or BBIBP-CorV (256 participants). All these participants that received the booster vaccination were included in the safety set (SS) for safety analysis. 255 participants in NVSI-06-09 booster group and 249 in BBIBP-CorV booster group who had valid immunogenicity data were included in the per-protocol set (PPS) for immunogenicity analysis.

Male participants accounted for 94.57% of the population, and Asian was the majority (93.99%). Baseline demographic characteristics were well-balanced between the NVSI-06-09 and BBIBP-CorV booster groups. The age composition, male-to-female ratio, race distribution and physiological status were quite similar across groups (all *p*>0.05) (Table 1). Before booster vaccination, 24 participants from NVSI-06-09 group and 23 from BBIBP-CorV group were tested positive or weak positive for COVID-19 infection by the swab PCR assay (Table 1).

**Table 1.** Demographic characteristics of the participants (FAS)

|  | **NVSI-06-09**<br>**(N=260)** | **BBIBP-CorV**<br>**(N=256)** | ***p* value** |
| --- | --- | --- | --- |
| Age (years) |  |  |  |
| Mean (SD) | 37.2 (9.1) | 37.3 (8.7) | 0.8740 |
| Median | 36.0 | 36.5 |  |
| Min, Max | 19, 62 | 19, 63 |  |
| Sex, n (%) |  |  | 0.4622 |
| Male | 244 (93.85) | 244 (95.31) |  |
| Female | 16 (6.15) | 12 (4.69) |  |
| Race |  |  | 0.4216 |
| Chinese | 0 (0.00) | 0 (0.00) |  |
| Asian (except Chinese) | 243 (93.46) | 242 (94.53) |  |
| Black/African/Caribbean | 6 (2.31) | 8 (3.13) |  |
| White | 11 (4.23) | 6 (2.34) |  |
| Multiracial | 0 (0.00) | 0 (0.00) |  |
| Other | 0 (0.00) | 0 (0.00) |  |
| Body temperature before<br>immunization (°C) |  |  |  |
| Mean (SD) | 36.41 (0.19) | 36.42 (0.20) | 0.5994 |
| Median | 36.40 | 36.40 |  |
| Min, Max | 36.0, 37.0 | 36.1, 37.0 |  |
| Height (cm) |  |  |  |
| Mean (SD) | 171.2 (7.2) | 170.9 (7.3) | 0.5493 |
| Median | 171.0 | 171.0 |  |
| Min, Max | 151, 195 | 146, 192 |  |
| Weight (kg) |  |  |  |
| Mean (SD) | 78.69 (14.52) | 80.96 (14.34) | 0.0745 |
| Median | 79.00 | 79.00 |  |
| Min, Max | 49.0, 131.0 | 49.0, 136.0 |  |
| Cardiopulmonary<br>auscultation |  |  | 1.0000 |
| Normal | 260 (100.00) | 256 (100.00) |  |
| Abnormal | 0 (0.00) | 0 (0.00) |  |
| PCR test before<br>immunization, n (%) |  |  | 0.8935 |
| Negative | 236 (90.77) | 233 (91.02) |  |
| Positive | 11 (4.23) | 9 (3.52) |  |
| Weak positive | 13 (5.00) | 14 (5.47) |  |

The *p* value for comparing the NVSI-06-09 and BBIBP-CorV groups were calculated by using two-sided student's t-test for continuous data (after log-transformation of antibodies) and two-sided Chi-square test for non-ordered categorical data.
n = participant number.
SD = standard deviation, PCR = polymerase Chain Reaction.

### SAFETY

At the time of writing this report, no serious adverse event or adverse event of special interest was observed. Within 7 days after booster vaccination, 31 participants (11.92%) in NVSI-06-09 group and 34 (13.28%) in BBIBP-CorV group reported at least one solicited adverse reaction, and all of them were grade 1 or 2. The overall incidence of solicited adverse reactions was low and similar between two groups (*p*=0.6915) (Fig. 2 and Table S1). The local adverse reactions reported were injection site pain of grade 1 or 2 by NVSI-06-09 group, and injection site pain (grade 1) and pruritus (grade 1) by BBIBP-CorV group. The systemic adverse reactions reported by both groups were mainly muscle pain (non-vaccination site), headache, fever, and fatigue (Fig. 2 and Table S1). Within 30 days post-booster, 8 (3.08%) participants in NVSI-06-09 group and 7 (2.73%) in BBIBP-CorV group reported unsolicited adverse reactions, all of which were grade 1 or 2. The incidence and severity of unsolicited adverse reactions were low and similar between two booster groups (*p*=1.0000) (Table S1). Overall, both the NVSI-06-09 and BBIBP-CorV booster vaccinations showed good tolerance and safety with limited adverse reactions observed, and the safety profile was comparable between two groups.

**Figure 2.**
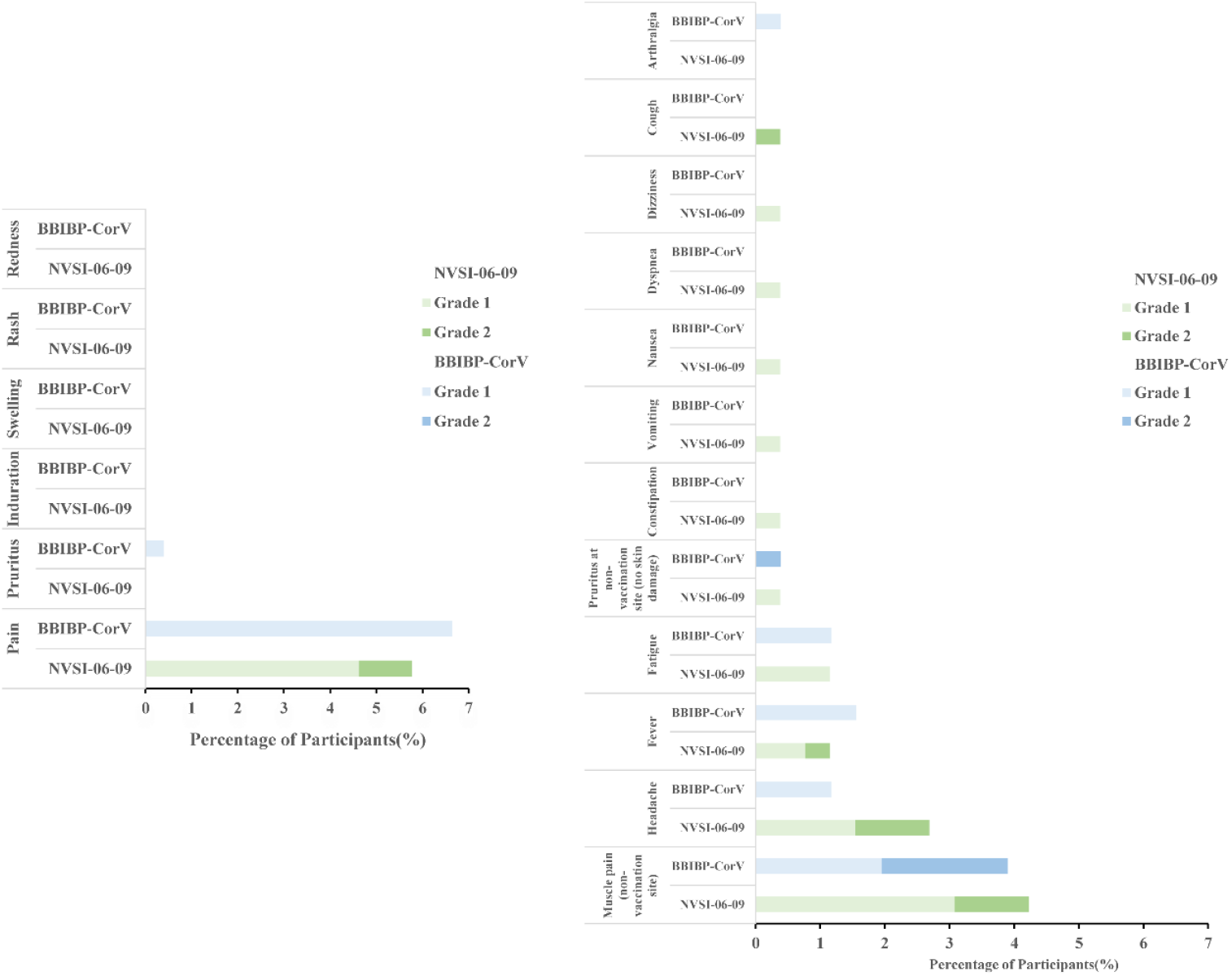
The incidence and severity of adverse reactions in NVSI-06-09 booster group compared with those in BBIBP-CorV booster group. (A) Local adverse reactions reported within 7 days after the administration of booster vaccination. (B) Systemic reactions recorded within 7 days after booster vaccination. Adverse reactions are graded according to the relevant guidance of China National Medical Products Administration (NMPA).

### IMMUNOGENICITY AGAINST PROTOTYPE SARS-COV-2 VIRUS AND OMICRON BA.1.1 VARIANT

The immunogenicity of NVSI-06-09 as a booster dose was evaluated and compared with BBIBP-CorV by using live-virus microneutralization assay. Before booster vaccination, neutralizing antibody (nAb) against the prototype SARS-CoV-2 virus was detectable (nAb titer ≥4) in most participants in NVSI-06-09 (99.61%) and BBIBP-CorV (99.20%) groups, with nAb geometric mean titers (GMTs) of 119.42 (95%CI, 106.47-133.95) and 113.29 (100.50-127.71), respectively (Fig. 3 and Table S2). After booster vaccination, nAb response against the prototype virus was significantly improved in both groups, but the post-booster antibody titers induced by NVSI-06-09 booster were distinctly higher than those boosted by BBIBP-CorV. On day 14 post-booster, nAb GMT against the prototype stain in the homologous BBIBP-CorV booster group increased to 506.47 (95% CI, 458.25-559.77), with a 4.47-fold (4.01-4.99) rise compared to the pre-booster baseline. Whereas, neutralizing GMT in the NVSI-06-09 booster group reached 2349.44 (95% CI, 2061.42-2677.70), with a remarkably 19.67-fold (16.79-23.06) increase from baseline. Correspondingly, the four-fold rise rate of nAb titers against the prototype virus induced by NVSI-06-09 was 93.33% (95% CI, 89.54%-96.07%), which was much greater than 59.44% (53.06%-65.59%) by BBIBP-CorV (Fig. 3 and Table S2). Some participants exhibited positive (or weak positive) COVID-19 PCR tests before booster vaccination. After excluding these positive participants, similar results were also obtained (Table S2). Then, we compared the neutralizing responses between two booster groups stratified by different doses (i.e., two or three doses) of prior primary BBIBP-CorV vaccinations. For the participants receiving two-dose primary vaccinations, the fold rise of nAb GMT induced by the booster of NVSI-06-09 was significantly greater than that of the BBIBP-CorV booster (15.26-fold vs 2.77-fold) (Table S2). A similar trend was also observed in the participants with three-dose primary vaccinations, where the GMT fold rise was distinctly higher in NVSI-06-09 group than BBIBP-CorV group (19.90-fold vs 4.55-fold) (Table S2).

**Figure 3.**
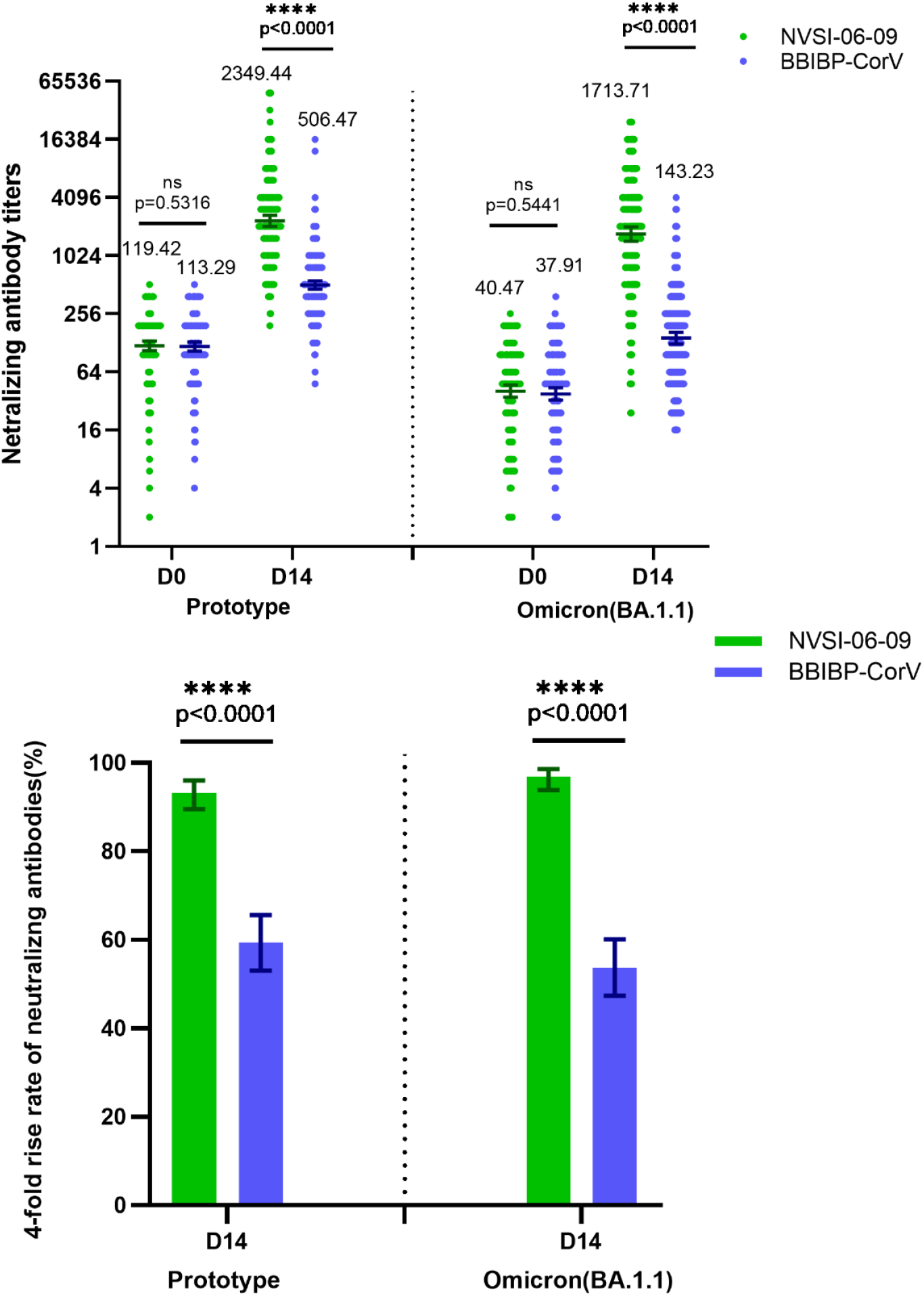
The live-virus neutralizing antibody titers, along with the corresponding four-fold rise rate, against prototype SARS-CoV-2 strain and Omicron BA.1.1 variant induced by the booster vaccination of NVSI-06-09, compared with those boosted with BBIBP-CorV. (A) The anti-prototype and anti-Omicron BA.1.1 neutralizing antibody titers at baseline and on day 14 after booster vaccination. Data are presented as GMTs and 95% CIs. (B) The corresponding four-fold rise rate of neutralizing antibodies from baseline and 95% CIs on day 14 after booster. ****: *p*<0.0001, ns: not significant.

At pre-booster baseline, 96.86% participants in NVSI-06-09 group and 97.19% in BBIBP-CorV group exhibited detectable nAbs against the Omicron BA.1.1 variant, with the GMT values of 40.47 (95% CI, 34.86-46.97) and 37.91 (32.64-44.04), respectively (Fig. 3 and Table S3). The baseline level of nAb GMT against Omicron BA.1.1 variant was obviously lower than that against the prototype strain, indicating that the Omicron BA.1.1 variant was partially resistant to two- or three-dose BBIBP-CorV vaccinations. Both NVSI-06-09 and BBIBP-CorV booster vaccinations markedly elevated Omicron BA.1.1-specific nAb response, and similar to that of prototype virus, the nAb level against Omicron BA.1.1 variant elicited by NVSI-06-09 was dramatically higher than that induced by BBIBP-CorV. On day 14 post-booster, the increase of Omicron BA.1.1-specific nAbs from pre-booster baseline by NVSI-06-09 arrived at 42.35-fold (95% CI, 35.15-51.03) with GMT of 1713.71 (95% CI, 1447.12-2029.41), and 3.78-fold (3.40-4.20) by BBIBP-CorV with GMT of 143.23 (124.48-164.80). Correspondingly, the four-fold rise rate of nAb titers against the Omicron BA.1.1 induced by NVSI-06-09 booster was also significantly greater than that by BBIBP-CorV booster (96.86% vs 53.82%) (Fig. 3 and Table S3). After excluding the participants with pre-booster positive COVID-19 infections, similar results were also obtained (Table S3). When investigating the participants separately with two or three primary doses, similar superiority was observed in NVSI-06-09 group. The NVSI-06-09 booster elicited 27.48-fold (95% CI, 7.32-103.15) and 43.18-fold (35.78-52.11) rises of Omicron BA.1.1-specific nAb GMT in participants receiving two and three primary doses, respectively. In contrast, the BBIBP-CorV booster only induced 1.49-fold (95% CI, 0.94-2.37) increases in two-dose regimen and 3.91-fold (3.51-4.35) in three-dose regimen (Table S3).

As demonstrated by the results presented above, BBIBP-CorV booster elicited similar or lower nAb GMT fold rise against Omicron BA.1.1 variant than that against prototype strain (3.78-fold vs 4.47-fold). By contrary, NVSI-06-09 booster induced a much greater GMT fold rise against Omicron BA.1.1 strain than that against prototype virus (42.35-fold vs 19.67-fold). These results indicated that NVSI-06-09 booster exhibited remarkably better immunogenicity against SARS-CoV-2 virus, especially Omicron BA.1.1 variant.

### CROSS-REACTIVE IMMUNOGENICITY AGAINST OTHER OMICRON SUB-VARIANTS

SARS-CoV-2 Omicron variant evolves rapidly, and many sub-variants have emerged. Owing to its higher transmissibility and immune-evasive capability, Omicron BA.2 replaced BA.1 quickly, and was soon replaced by BA.4 and BA.5 afterwards. Currently, BA.4 and BA.5 have become the dominantly circulating variant worldwide. Besides Omicron BA.1.1, the cross-reactive immunogenicity of NVSI-06-09 against BA.2, BA.4 and BA.5 was also evaluated by using the live-virus microneutralization assay. The nAb responses against these three Omicron sub-variants were tested in all participants in PPS, and compared between the NVSI-06-09 and BBIBP-CorV booster groups.

Before booster vaccination, 96.85%-97.64% participants in NVSI-06-09 group, and 95.58%-97.99% in BBIBP-CorV group have detectable nAbs against the Omicron BA.2, BA.4 and BA.5 sub-variants. The anti-BA.2, anti-BA.4 and anti-BA.5 neutralizing baseline GMTs were similar between two groups, i.e., 39.19 (95% CI, 34.27-44.82), 40.59 (35.75-46.10) and 36.95 (32.38-42.16) in the NVSI-06-09 booster group, and 40.35 (35.05-46.45), 42.14 (36.80-48.25) and 34.98 (30.53- 40.09) in the BBIBP-CorV booster group (Fig. 4, and Tables S4, S5 and S6). The pre-booster GMTs against three Omicron sub-variants were noticeably less than that against prototype virus, implying less sensitive of these Omicron sub-variants to primary BBIBP-CorV vaccinations. On day 14 after booster vaccination, the BA.2-, BA.4- and BA.5-specific neutralizing GMTs in BBIBP-CorV booster group increased to 117.33 (95% CI, 103.07-133.55), 113.51 (98.50-130.81) and 165.39 (142.99-191.30), with 2.91-fold (95% CI, 2.60-3.25), 2.69-fold (2.38-3.05) and 4.73-fold (4.10-5.45) increase from baselines, respectively. By contrast, a dramatically higher neutralizing response was induced by NVSI-06-09 booster, in which the BA.2-, BA.4- and BA.5-specific neutralizing GMTs reached 983.36 (95% CI, 852.25-1134.64), 910.19 (779.53-1062.75) and 999.85 (872.41-1145.90) with the GMT fold rises of 25.09-fold (95% CI, 21.17-29.73), 22.42-fold (18.78-26.77) and 27.06-fold (23.04-31.78), respectively (Fig. 4, and Tables S4, S5 and S6). The remarkably better immunogenicity of NVSI-06-09 can also be observed from the four-fold rise rates in nAb titers. In participants from NVSI-06-09 booster group, the four-fold rise rates of anti-BA.2, anti-BA.4 and anti-BA.5 neutralizing titers were 94.49% (95% CI, 90.92%-96.95%), 92.52 % (95% CI, 88.56%-95.44%) and 94.88 % (95% CI, 91.41%-97.25%), respectively, which were significantly higher than 41.77% (95% CI, 35.57%-48.16%), 42.97% (95% CI, 36.74%-49.37%) and 59.04% (95% CI, 52.65%-65.20%) in BBIBP-CorV booster group (Fig. 4, and Tables S4, S5 and S6). After excluding the participants with pre-booster positive COVID-19 infections, similar results were obtained (Tables S4, S5 and S6). Similar results were also obtained when participants were stratified by two-dose or three-dose primary vaccination regimens. Whether two or three doses of BBIBP-CorV were administered previously by the participants, a booster dose of NVSI-06-09 induce much higher nAb responses against Omicron BA.2, BA.4 and BA.5 sub-variants than those boosted by BBIBP-CorV (Tables S4, S5 and S6).

**Figure 4.**
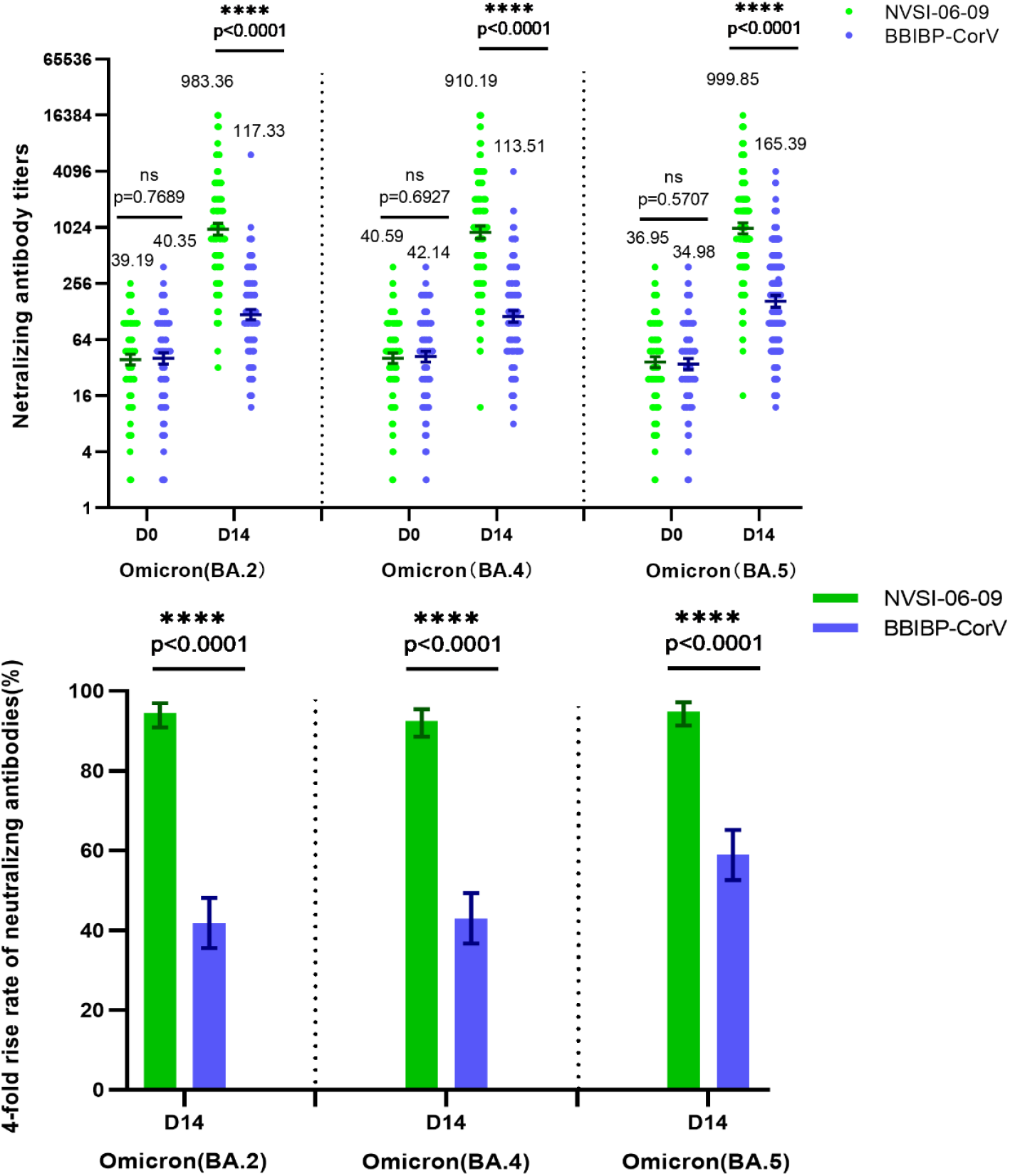
The cross-neutralizing antibody titers, along with the corresponding four-fold rise rate, against Omicron BA.2, BA.4 and BA.5 induced by the booster vaccination of NVSI-06- 09, compared with those boosted with BBIBP-CorV. (A) The anti-BA.2, anti-BA.4 and anti-BA.5 neutralizing antibody titers at baseline and on day 14 after booster vaccination. Data are presented as GMTs and 95% CIs. (B) The corresponding four-fold rise rate of neutralizing antibodies from baseline and 95%CIs on day 14 after booster. ****: *p*<0.0001, ns: not significant.

### CROSS-REACTIVE IMMUNOGENICITY AGAINST BETA AND DELTA VARIANTS

NVSI-06-09 is designed as a potential broad-spectrum COVID-19 vaccine. Besides the Omicron variants, the cross-reactive immunogenicity of NVSI-06-09 booster was also tested against other immune-evasive variants including Beta and Delta, using BBIBP-CorV as a comparison. A subset of serum samples with sequential numbers collected from 99 participants in NVSI-06-09 group and 100 participants in BBIBP-CorV group were used to evaluate the cross-neutralization against Beta and Delta variants.

The results showed that nAb GMTs against Beta and Delta variants induced by NVSI-06-09 booster were 3075.59 (95% CI, 2393.00-3952.89) and 2831.50 (2200.42-3643.58), respectively, which were 6.60-fold and 7.17-fold higher than those boosted by BBIBP-CorV with the GMT values of 465.69 (95% CI, 376.75-575.63) and 395.05 (316.82-492.58) (Fig. 5 and Table S7). In particular, the nAb GMTs induced by NVSI-06-09 booster against Beta and Delta variants were comparable to or even higher than those against the prototype strain.

**Figure 5.**
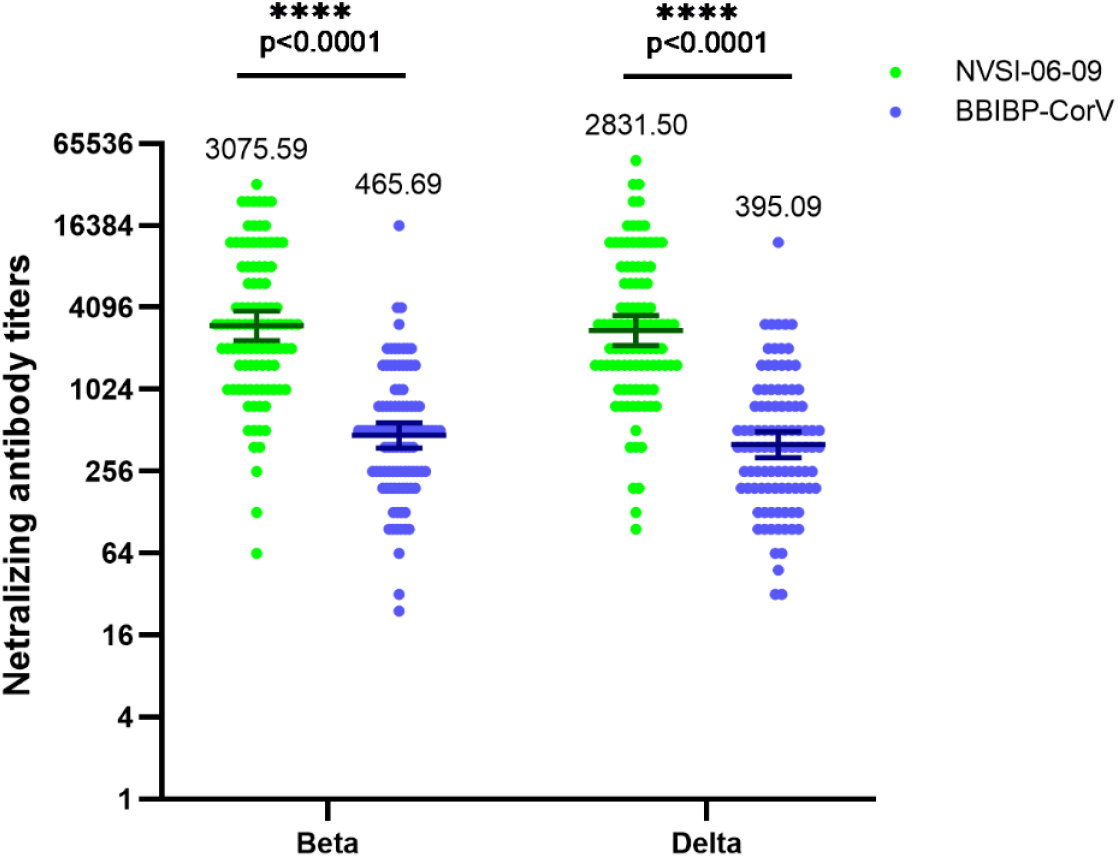
The cross-neutralizing antibody titers against Beta and Delta variants induced by the booster vaccination of NVSI-06-09, compared with those boosted with BBIBP-CorV. On 14 days after booster vaccination, a subset of 99 serum samples in NVSI-06-09 booster group and 100 samples in BBIBP-CorV booter group with sequential numbers were tested by using live-virus microneutralization assay. Data are presented as GMTs and 95% CIs. ****: *p*<0.0001.

## DISCUSSION

NVSI-06-09 is a potentially broad-spectrum vaccine using a mosaic-type trimeric RBD protein as the antigen, which combined key mutations from Omicron and other circulating variants into a single molecule. The interim results of the current study showed that a booster injection of NVSI-06-09 in participants who had completed two- or three-dose vaccinations of BBIBP-CorV was safe and well-tolerant. The safety profile of NVSI-06-09 booster was very similar to that of BBIBP-CorV booster, which was also comparable to the safety of two-dose primary vaccination with BBIBP-CorV as reported by Xia et al.^14^ Although most of participants in this trial had received three doses of BBIBP-CorV prior to enrollment, an additional booster dose of NVSI-06-09 or BBIBP-CorV did not bring additional safety risk of concern. For immunogenicity, NVSI-06-09 booster induced not only much higher but also much broader neutralizing responses than BBIBP-CorV booster against divergent SARS-CoV-2 stains. After booster vaccination, the nAb GMT against prototype virus in NVSI-06-09 group was 4.57-fold (after adjustment) higher than that in BBIBP-CorV group. Whereas, the ratios of neutralizing GMT after adjustment between two groups against Omicron BA.1.1, BA.2, BA.4 and BA.5 were improved to 11.61-fold, 8.50-fold, 8.15-fold, and 5.92-fold, and those against Beta and Delta variants were improved to 6.60-fold and 7.17-fold. It has been widely reported that nAb levels are highly correlated with immune protection against SARS-CoV-2.^15-17^ The stronger and broader nAb titers induced by NVSI-06-09 indicate that this vaccine may serve as a promising booster vaccine to better combat divergent SARS-CoV-2 variants, especially the immune-evasive variant of Omicron and its sub-lineages.

WHO has recommended a booster dose or even a second booster dose of COVID-19 vaccine to better combat the pandemic of SARS-CoV-2 (https://www.who.int/publications/). Safety of the vaccine is vital for the large-scale rollout of booster vaccinations. NVSI-06-09 is highly immunogenic and also has a good safety profile, supporting it as a promising booster candidate. Its high safety is attributed to the use of aluminum hydroxide as the adjuvant, whose safety has been extensively validated. Our data indicated that aluminum adjuvant combined with a well-designed antigen is a safe yet effective choice for booster vaccination in large-scale population.

As an advanced vaccine developed based on NVSI-06-07, our first-generation trimeric RBD-based vaccine targeting the prototype SARS-CoV-2 virus,^10,11^ NVSI-06-09 exhibited stronger immunogenic activities against the immune-evasive variants, including Omicron. Preclinical animal studies showed that NVSI-06-09 elicited higher cross-neutralization to Omicron and other circulating variants than NVSI-06-07.^12^ Comparing the results of this trial with the data from the trial on NVSI-06-07 reported previously,^11^ the nAb levels boosted by NVSI-06-09 were remarkably higher than those by NVSI-06-07 against SARS-CoV-2 variants, especially Omicron. It should be mentioned that most of the participants in this trial had received three doses of BBIBP-CorV before booster vaccination, and those in the trial for NVSI-06-07 received two doses. A thorough head-to-head comparison between NVSI-06-09 and NVSI-06-07 for the immunogenicity will be conducted in the future.

To date, COVID-19 vaccines authorized by WHO for emergency use are all targeting the prototype SARS-CoV-2 virus. Numerous studies have shown that after the primary-series vaccination of prototype vaccines, the nAb titers against Beta, Delta and Omicron variants decreased by 1.07-16.32-fold, 1.0-7.2-fold and 1.15-109.87-fold, respectively, when compared with that against the prototype virus.^18^ Although booster vaccinations could significantly improve the immune responses, the neutralization against Beta, Delta and Omicron variants was still 0.86-15-fold, 0.76-9.23-fold and 1.15-26.87-fold, respectively, lower than that against the prototype strain.^18^ Even within the Omicron variant, BA.4/BA.5 sub-variants also largely evaded the neutralization induced by BA.1 infection or vaccination, with a 2.6-8.0-fold drop in the nAb titers.^19-21^ Therefore, there is an urgent need to develop broad-spectrum vaccines to fight against immune-evasive variants. The findings from our study demonstrated that a booster shot of NVSI-06-09 not only significantly improved but also broadened the immune response to SARS-CoV-2 variants, which may serve as a candidate vaccine with potentially broad-spectrum immune activity.

Aiming at the defending against the emergence of immune-evasive variants of SARS-CoV-2, several variant-specific vaccines have been developed. Utilizing the inactivated virus-based, mRNA-based, and recombinant protein-based platforms, multiple updated vaccines specific to Beta, Delta, Omicron BA.1 or Omicron BA.2 have been developed.^20,22-28^ Results of animal studies and clinical trials have shown that these variant-targeting vaccines could induce higher nAb responses than the ancestral vaccine against these specific variants. Furthermore, variant-specific vaccines along with prototype vaccines have been used in combination to achieve cross-neutralizing responses against divergent SARS-CoV-2 strains (ClinicalTrials.gov identifier: NCT05365724, NCT05381350, NCT05382871).^20,23,26,28^ We, alternatively, adopt a new strategy on the principle of mosaic-type antigen for vaccine development to enable broad neutralization against SARS-CoV-2 variants. This strategy has been successfully applied in development of broad-spectrum vaccine against other highly mutated viruses, such as HIV and influenza,^29-33^ and several studies indicated that the mosaic antigen could elicit broader antibody responses than those induced by the mixture of the corresponding monovalent antigens.^31^ Our trial results demonstrated that the development of mosaic-type vaccines may provide a promising approach to better control the pandemic of the existing and future SARS-CoV-2 variants.

The uncertainty of SARS-CoV-2 evolution may further lead to increased transmissibility, immune evasion properties, and virulence of the virus in the future. It is necessary to accumulate effective technologies to better cope with new variants. The construction of mosaic vaccine may serve as one of the effective strategies against divergent SARS-CoV-2 variants.

Our study has some limitations. Firstly, only a small percentage of participants who received two doses of BBIBP-CorV were enrolled in the study. The potential influence of primary dosing frequency of BBIBP-CorV on the immune response boosted by NVSI-06-09 may not be fully revealed. Secondly, very few female volunteers participated in the trial, and thus the results obtained may not well reflect the potential immune response in females.

## Data Availability

The clinical trial is still ongoing, and the data will be available when the trial is complete upon request to the corresponding author (Q.M.L.). After study proposals are approved, data can be shared through secure online platforms.

## AUTHOR CONTRIBUTIONS

N.A.K. was the chief investigator. N.A.K., Q.M.L., Y.T.Z., X.M.Y., Y.K.Y., JingZ.(NVSI) and Y.K. designed the trial and study protocol. Q.M.L., JingZ.(NVSI), Y.L. and J.G.S. designed the recombinant vaccine NVSI-06-09. Q.M.L., Y.T.Z. and X.M.Y. were responsible for the organization and supervision of the project. X.F.Z., Y.Q.J., H.Z., Z.M.L., Y.N.H., J.W.H., S.C., Z.J.M., X.J.G. and X.Y.M. contributed to the production of NVSI-06-09 vaccine. H.W. and JinZ. provided the inactivated vaccine BBIBP-CorV. X.M.Y., Y.T.Z., I.E. and Y.K.Y. contributed to project management. T.Y., Y.K., M.L., L.Q., W.Z., P.X., X.Z., C.Q., D.Y.L. and S.S.Y. participated in the implementation of the trial. H.M.M., Z.N.W. and J.L.Y. conducted sample collection and processing. G.W., K.X., W.L., JingZ.(CDC), M.Y. and S.M. carried out the serum tests. G.W., K.X., Y.L. N.L., X.Z, F.J.S., Z.B.H., F.T., F.Z., J.J.W., X.Y.L, X.L., L.F.D. and Z.H.L. contributed to the development of serum testing method and manuscript preparation. S.H. and M.S.E. performed the clinical data gathering, analysis and operation. Z.J., X.C. and Y.T. performed statistical analysis of the data. Q.M.L., JingZ.(NVSI), Y.L., G.W., Y.K.Y. and K.X. analyzed and interpreted the results. J.G.S., Y.L., JingZ.(NVSI), K.X., L.F.D., S.S. and S.S.Y. contributed to the writing of the manuscript. Q.M.L. and G.W. revised the manuscript. All authors approved the final manuscript.

## ACKNOWLEDGEMENTS

The study was sponsored by China National Biotec Group Co., Ltd. (CNBG) of Sinopharm, National Vaccine and Serum Institute (NVSI) of Sinopharm CNBG, Lanzhou Institute of Biological Products Co., Ltd. (LIBP) of Sinopharm CNBG, and Beijing Institute of Biological Products Co., Ltd. (BIBP) of Sinopharm CNBG.

---

**Table S1.** Solicited and unsolicited adverse reactions after booster vaccination.

|  | **NVSI-06-09**<br>**(N=260)** | **BBIBP-CorV**<br>**(N=256)** | **Total**<br>**(N=516)** | ***p* value\*** |
| --- | --- | --- | --- | --- |
| **Solicited adverse reactions within 0-7 days** | 31(11.92) | 34(13.28) | 65(12.60) | 0.6915 |
| Grade 1 | 26(10.00) | 30(11.72) | 56(10.85) | 0.5728 |
| Grade 2 | 6(2.31) | 6(2.34) | 12(2.33) | 1.0000 |
| **Injection site adverse reactions** | 15(5.77) | 18(7.03) | 33(6.40) | 0.5932 |
| Grade 1 | 12(4.62) | 18(7.03) | 30(5.81) | 0.2636 |
| Grade 2 | 3(1.15) | 0(0.00) | 3(0.58) | 0.2486 |
| Pain | 15(5.77) | 17(6.64) | 32(6.20) | 0.7181 |
| Grade 1 | 12(4.62) | 17(6.64) | 29(5.62) | 0.3445 |
| Grade 2 | 3(1.15) | 0(0.00) | 3(0.58) | 0.2486 |
| Pruritus | 0(0.00) | 1(0.39) | 1(0.19) | 0.4961 |
| Grade 1 | 0(0.00) | 1(0.39) | 1(0.19) | 0.4961 |
| Induration | 0(0.00) | 0(0.00) | 0(0.00) | 1.0000 |
| Swelling | 0(0.00) | 0(0.00) | 0(0.00) | 1.0000 |
| Rash | 0(0.00) | 0(0.00) | 0(0.00) | 1.0000 |
| Redness | 0(0.00) | 0(0.00) | 0(0.00) | 1.0000 |
| **Systemic adverse reactions** | 20(7.69) | 19(7.42) | 39(7.56) | 1.0000 |
| Grade 1 | 16(6.15) | 14(5.47) | 30(5.81) | 0.8513 |
| Grade 2 | 5(1.92) | 6(2.34) | 11(2.13) | 0.7705 |
| Muscle pain (non-vaccination site) | 11(4.23) | 10(3.91) | 21(4.07) | 1.0000 |
| Grade 1 | 8(3.08) | 5(1.95) | 13(2.52) | 0.5763 |
| Grade 2 | 3(1.15) | 5(1.95) | 8(1.55) | 0.5015 |
| Headache | 7(2.69) | 3(1.17) | 10(1.94) | 0.3391 |
| Grade 1 | 4(1.54) | 3(1.17) | 7(1.36) | 1.0000 |
| Grade 2 | 3(1.15) | 0(0.00) | 3(0.58) | 0.2486 |
| Fever | 3(1.15) | 4(1.56) | 7(1.36) | 0.7229 |
| Grade 1 | 2(0.77) | 4(1.56) | 6(1.16) | 0.4472 |
| Grade 2 | 1(0.38) | 0(0.00) | 1(0.19) | 1.0000 |
| Fatigue | 3(1.15) | 3(1.17) | 6(1.16) | 1.0000 |
| Grade 1 | 3(1.15) | 3(1.17) | 6(1.16) | 1.0000 |
| Pruritus at non-vaccination site (no skin damage) | 1(0.38) | 1(0.39) | 2(0.39) | 1.0000 |
| Grade 1 | 1(0.38) | 0(0.00) | 1(0.19) | 1.0000 |
| Grade 2 | 0(0.00) | 1(0.39) | 1(0.19) | 0.4961 |
| Arthralgia | 0(0.00) | 1(0.39) | 1(0.19) | 0.4961 |
| Grade 1 | 0(0.00) | 1(0.39) | 1(0.19) | 0.4961 |
| Constipation | 1(0.38) | 0(0.00) | 1(0.19) | 1.0000 |
| Grade 1 | 1(0.38) | 0(0.00) | 1(0.19) | 1.0000 |
| Vomiting | 1(0.38) | 0(0.00) | 1(0.19) | 1.0000 |
| Grade 1 | 1(0.38) | 0(0.00) | 1(0.19) | 1.0000 |
| Nausea | 1(0.38) | 0(0.00) | 1(0.19) | 1.0000 |
| Grade 1 | 1(0.38) | 0(0.00) | 1(0.19) | 1.0000 |
| Cough | 1(0.38) | 0(0.00) | 1(0.19) | 1.0000 |
| Grade 1 | 0(0.00) | 0(0.00) | 1(0.19) | 1.0000 |
| Grade 2 | 1(0.38) | 0(0.00) | 1(0.19) | 1.0000 |
| Dyspnea | 1(0.38) | 0(0.00) | 1(0.19) | 1.0000 |
| Grade 1 | 1(0.38) | 0(0.00) | 1(0.19) | 1.0000 |
| Dizziness | 1(0.38) | 0(0.00) | 1(0.19) | 1.0000 |
| Grade 1 | 1(0.38) | 0(0.00) | 1(0.19) | 1.0000 |
| Diarrhea | 0(0.00) | 0(0.00) | 0(0.00) | 1.0000 |
| Dysphagia | 0(0.00) | 0(0.00) | 0(0.00) | 1.0000 |
| Anorexia | 0(0.00) | 0(0.00) | 0(0.00) | 1.0000 |
| Abnormal skin mucosa | 0(0.00) | 0(0.00) | 0(0.00) | 1.0000 |
| Acute allergic reaction | 0(0.00) | 0(0.00) | 0(0.00) | 1.0000 |
| **Unsolicited adverse reactions within 0-30 days** | 8(3.08) | 7(2.73) | 15(2.91) | 1.0000 |
| Grade 1 | 7(2.69) | 3(1.17) | 10(1.94) | 0.3391 |
| Grade 2 | 1(0.38) | 4(1.56) | 5(0.97) | 0.2133 |

Notes: \**p*-value was calculated using Fisher's exact test. Data are presented as n (%).

**Table S2.** Live-virus neutralizing antibody response against prototype SARS-CoV-2 strain (PPS)

|  | 14 days after boosting |  |  |
| --- | --- | --- | --- |
|  | NVSI-06-09 | BBIBP-CorV | *p* value |
| **All subjects** |  |  |  |
| N(missing) | 255(0) | 249(0) |  |
| Pre-booster antibody titer $\geq 4$, n (%) | 254(99.61) | 247(99.20) | 0.6198 |
| 95% CI (%) | 97.83-99.99 | 97.13-99.90 |  |
| Pre-booster antibody GMT<sup>[1]</sup> (95%CI) | 119.42(106.47-133.95) | 113.29(100.50-127.71) | 0.5316 |
| Post-booster antibody GMT (95%CI) | 2349.44(2061.42-2677.70) | 506.47(458.25-559.77) | <0.0001 |
| Ratio of GMT between two groups(95%CI)<sup>[2]</sup> | 4.64(3.93-5.47) |  |  |
| Post-booster adjusted antibody GMT (95%CI)<sup>[3]</sup> | 1982.55(1601.51-2454.25) | 433.41(349.04-538.18) | <0.0001 |
| Ratio of adjusted GMT between two groups(95%CI)<sup>[3]</sup> | 4.57(3.91-5.35) |  |  |
| Rate of 4-fold rise<sup>[4]</sup>, n (%) | 238(93.33) | 148(59.44) | <0.0001 |
| 95% CI (%) | 89.54-96.07 | 53.06-65.59 |  |
| Rate difference between two groups (% , 95%CI<sup>[5]</sup>) | 33.95(27.13-40.77) |  |  |
| Post-booster antibody GMT fold rise (95%CI) | 19.67(16.79-23.06) | 4.47(4.01-4.99) | <0.0001 |
| **All subjects (Excluding participants with positive or weak positive COVID-19 PCR test)** |  |  |  |
| N(missing) | 231(0) | 226(0) |  |
| Pre-booster antibody titer $\geq 4$, n (%) | 230(99.57) | 225(99.56) | 1.0000 |
| 95% CI (%) | 97.61-99.99 | 97.56-99.99 |  |
| Pre-booster antibody GMT<sup>[1]</sup> (95%CI) | 120.32(106.60-135.80) | 115.90(102.63-130.88) | 0.6675 |
| Post-booster antibody GMT (95%CI) | 2321.93(2020.94-2667.74) | 514.35(462.45-572.08) | <0.0001 |
| Ratio of GMT between two groups(95%CI)<sup>[2]</sup> | 4.51(3.79-5.38) |  |  |
| Post-booster adjusted antibody GMT (95%CI)<sup>[3]</sup> | 1867.43(1490.04-2340.41) | 415.29(329.12-524.02) | <0.0001 |
| Ratio of adjusted GMT between two groups(95%CI)<sup>[3]</sup> | 4.50(3.81-5.31) |  |  |
|  | NVSI-06-09 | BBIBP-CorV | *p* value |
| Rate of 4-fold rise<sup>[4]</sup>, n (%) | 214(92.64) | 134(59.29) | <0.0001 |
| 95% CI (%) | 88.48-95.65 | 52.58-65.76 |  |
| Rate difference between two groups (% , 95% CI<sup>[5]</sup>) | 33.57(26.34-40.80) |  |  |
| Post-booster antibody GMT fold rise (95% CI) | 19.30(16.31-22.83) | 4.44(3.96-4.98) | <0.0001 |
| **Subjects who have received 2 doses of inactivated vaccine previously** |  |  |  |
| N(missing) | 11(0) | 9(0) |  |
| Pre-booster antibody titer $\geq 4$, n (%) | 11(100.00) | 9(100.00) | 1.0000 |
| 95% CI (%) | 71.51-100.00 | 66.37-100.00 |  |
| Pre-booster antibody GMT<sup>[1]</sup> (95% CI) | 83.73(44.50-157.55) | 183.54(119.77-281.26) | 0.0410 |
| Post-booster antibody GMT (95% CI) | 1277.47(682.73-2390.30) | 509.04(298.48-868.14) | 0.0247 |
| Ratio of GMT between two groups(95% CI)<sup>[2]</sup> | 2.51(1.14-5.52) |  |  |
| Post-booster adjusted antibody GMT (95% CI)<sup>[3]</sup> | 1271.04(712.48-2267.47) | 512.19(268.04-978.74) | 0.0521 |
| Ratio of adjusted GMT between two groups(95% CI)<sup>[3]</sup> | 2.48(0.99-6.22) |  |  |
| Rate of 4-fold rise<sup>[4]</sup>, n (%) | 10(90.91) | 4(44.44) | 0.0279 |
| 95% CI (%) | 58.72-99.77 | 13.70-78.80 |  |
| Rate difference between two groups (% , 95% CI<sup>[5]</sup>) | 46.46(9.82-83.10) |  |  |
| Post-booster antibody GMT fold rise (95% CI) | 15.26(5.84-39.85) | 2.77(1.64-4.70) | 0.0042 |
| **Subjects who have received 3 doses of inactivated vaccine previously** |  |  |  |
| N(missing) | 244(0) | 240(0) |  |
| Pre-booster antibody titer $\geq 4$, n (%) | 243(99.59) | 238(99.17) | 0.6211 |
| 95% CI (%) | 97.74-99.99 | 97.02-99.90 |  |
| Pre-booster antibody GMT<sup>[1]</sup> (95% CI) | 121.35(107.93-136.44) | 111.26(98.38-125.82) | 0.3145 |
| Post-booster antibody GMT (95% CI) | 2414.87(2112.91-2759.98) | 506.38(457.01-561.08) | <0.0001 |
| Ratio of GMT between two groups(95% CI)<sup>[2]</sup> | 4.77(4.03-5.64) |  |  |
|  | NVSI-06-09 | BBIBP-CorV | *p* value |
| Post-booster adjusted antibody GMT (95%CI)<sup>[3]</sup> | 2382.06(2128.58-2665.72) | 513.47(458.40-575.15) | <0.0001 |
| Ratio of adjusted GMT between two groups(95%CI)<sup>[3]</sup> | 4.64(3.95-5.44) |  |  |
| Rate of 4-fold rise<sup>[4]</sup>, n (%) | 228(93.44) | 144(60.00) | <0.0001 |
| 95%CI (%) | 89.57-96.21 | 53.50-66.25 |  |
| Rate difference between two groups (% , 95%CI<sup>[5]</sup>) | 33.44(26.51-40.38) |  |  |
| Post-booster antibody GMT fold rise (95%CI) | 19.90(16.93-23.39) | 4.55(4.07-5.09) | <0.0001 |

Notes: [1] GMT represent geometric mean titer.
[2] The ratio of GMT between two groups was calculated by “NVSI-06-09/BBIBP-CorV”.
[3] Covariance analysis with least square method was used to calculate the adjusted GMT and the corresponding *p* value.
[4] Rate of 4-fold rise was defined as percentage of participants with a $\geq 4$ -fold rise in neutralizing antibody titer from baseline.
[5] Rate difference=(NVSI-06-09)-(BBIBP-CorV). Rate difference and 95%CI were estimated by CMH method considering stratification factors.

**Table S3.** Live-virus neutralizing antibody response against Omicron (BA.1.1) variant (PPS)

|  | 14 days after boosting |  |  |
| --- | --- | --- | --- |
|  | NVSI-06-09 | BBIBP-CorV | *p* value |
| **All subjects** |  |  |  |
| N(missing) | 255(0) | 249(0) |  |
| Pre-booster antibody titer $\geq 4$, n (%) | 247(96.86) | 242(97.19) | 0.8295 |
| 95% CI (%) | 93.91-98.64 | 94.29-98.86 |  |
| Pre-booster antibody GMT<sup>[1]</sup> (95%CI) | 40.47(34.86-46.97) | 37.91(32.64-44.04) | 0.5441 |
| Post-booster antibody GMT (95%CI) | 1713.71(1447.12-2029.41) | 143.23(124.48-164.80) | <0.0001 |
| Ratio of GMT between two groups(95%CI)<sup>[2]</sup> | 11.96(9.60-14.90) |  |  |
| Post-booster adjusted antibody GMT (95%CI)<sup>[3]</sup> | 1321.99(1019.80-1713.73) | 113.91(87.56-148.18) | <0.0001 |
| Ratio of adjusted GMT between two groups(95%CI)<sup>[3]</sup> | 11.61(9.60-14.03) |  |  |
| Rate of 4-fold rise<sup>[4]</sup>, n (%) | 247(96.86) | 134(53.82) | <0.0001 |
| 95% CI (%) | 93.91-98.64 | 47.41-60.13 |  |
| Rate difference between two groups (% , 95%CI<sup>[5]</sup>) | 43.21(36.69-49.73) |  |  |
| Post-booster antibody GMT fold rise (95%CI) | 42.35(35.15-51.03) | 3.78(3.40-4.20) | <0.0001 |
| **All subjects (Excluding participants with positive or weak positive COVID-19 PCR test)** |  |  |  |
| N(missing) | 231(0) | 226(0) |  |
| Pre-booster antibody titer $\geq 4$, n (%) | 224(96.97) | 220(97.35) | 0.8093 |
| 95% CI (%) | 93.86-98.77 | 94.31-99.02 |  |
| Pre-booster antibody GMT<sup>[1]</sup> (95%CI) | 41.67(35.69-48.65) | 38.41(32.81-44.97) | 0.4685 |
| Post-booster antibody GMT (95%CI) | 1732.14(1449.09-2070.47) | 149.81(129.37-173.48) | <0.0001 |
| Ratio of GMT between two groups(95%CI)<sup>[2]</sup> | 11.56(9.18-14.57) |  |  |
| Post-booster adjusted antibody GMT (95%CI)<sup>[3]</sup> | 1319.09(1003.59-1733.77) | 117.89(88.98-156.18) | <0.0001 |
| Ratio of adjusted GMT between two groups(95%CI)<sup>[3]</sup> | 11.19(9.15-13.68) |  |  |
|  | NVSI-06-09 | BBIBP-CorV | *p* value |
| Rate of 4-fold rise<sup>[4]</sup>, n (%) | 223(96.54) | 124(54.87) | <0.0001 |
| 95% CI (%) | 93.29-98.49 | 48.13-61.47 |  |
| Rate difference between two groups (% , 95% CI<sup>[5]</sup>) | 42.10(35.25-48.95) |  |  |
| Post-booster antibody GMT fold rise (95% CI) | 41.57(34.10-50.67) | 3.90(3.49-4.36) | <0.0001 |
| **Subjects who have received 2 doses of inactivated vaccine previously** |  |  |  |
| N(missing) | 11(0) | 9(0) |  |
| Pre-booster antibody titer ≥4, n (%) | 11(100.00) | 9(100.00) | 1.0000 |
| 95% CI (%) | 71.51-100.00 | 66.37-100.00 |  |
| Pre-booster antibody GMT<sup>[1]</sup> (95% CI) | 41.42(18.48-92.84) | 73.26(43.71-122.78) | 0.2212 |
| Post-booster antibody GMT (95% CI) | 1138.33(502.74-2577.48) | 109.09(53.40-222.87) | 0.0002 |
| Ratio of GMT between two groups(95% CI)<sup>[2]</sup> | 10.43(3.70-29.43) |  |  |
| Post-booster adjusted antibody GMT (95% CI)<sup>[3]</sup> | 1122.91(539.91-2335.46) | 110.93(49.20-250.11) | 0.0004 |
| Ratio of adjusted GMT between two groups(95% CI)<sup>[3]</sup> | 10.12(3.31-30.93) |  |  |
| Rate of 4-fold rise<sup>[4]</sup>, n (%) | 10(90.91) | 1(11.11) | 0.0005 |
| 95% CI (%) | 58.72-99.77 | 0.28-48.25 |  |
| Rate difference between two groups (% , 95% CI<sup>[5]</sup>) | 79.80(53.15-100.00) |  |  |
| Post-booster antibody GMT fold rise (95% CI) | 27.48(7.32-103.15) | 1.49(0.94-2.37) | 0.0005 |
| **Subjects who have received 3 doses of inactivated vaccine previously** |  |  |  |
| N(missing) | 244(0) | 240(0) |  |
| Pre-booster antibody titer ≥4, n (%) | 236(96.72) | 233(97.08) | 0.8183 |
| 95% CI (%) | 93.64-98.57 | 94.08-98.82 |  |
| Pre-booster antibody GMT<sup>[1]</sup> (95% CI) | 40.42(34.69-47.10) | 36.99(31.72-43.14) | 0.4202 |
| Post-booster antibody GMT (95% CI) | 1745.61(1467.36-2076.62) | 144.70(125.32-167.08) | <0.0001 |
| Ratio of GMT between two groups(95% CI)<sup>[2]</sup> | 12.06(9.63-15.11) |  |  |
|  | NVSI-06-09 | BBIBP-CorV | *p* value |
| Post-booster adjusted antibody GMT (95%CI)<sup>[3]</sup> | 1704.55(1488.14-1952.43) | 148.24(129.28-169.99) | <0.0001 |
| Ratio of adjusted GMT between two groups(95%CI)<sup>[3]</sup> | 11.50(9.48-13.94) |  |  |
| Rate of 4-fold rise<sup>[4]</sup>, n (%) | 237(97.13) | 133(55.42) | <0.0001 |
| 95%CI (%) | 94.18-98.84 | 48.89-61.81 |  |
| Rate difference between two groups (% , 95%CI<sup>[5]</sup>) | 41.71(35.09-48.34) |  |  |
| Post-booster antibody GMT fold rise (95%CI) | 43.18(35.78-52.11) | 3.91(3.51-4.35) | <0.0001 |

Notes: [1] GMT represent geometric mean titer.
[2] The ratio of GMT between two groups was calculated by “NVSI-06-09/ BBIBP-CorV”.
[3] Covariance analysis with least square method was used to calculate the adjusted GMT and the corresponding *p* value.
[4] Rate of 4-fold rise was defined as percentage of participants with a $\geq 4$ -fold rise in neutralizing antibody titer from baseline.
[5] Rate difference=(NVSI-06-09)-(BBIBP-CorV). Rate difference and 95%CI were estimated by CMH method considering stratification factors.

**Table S4.** Live-virus neutralizing antibody response against Omicron (BA.2) variant (PPS)

|  | 14 days after boosting |  |  |
| --- | --- | --- | --- |
|  | NVSI-06-09 | BBIBP-CorV | *p* value |
| **All subjects** |  |  |  |
| N(missing) | 255(1)<sup>[6]</sup> | 249(0) |  |
| Pre-booster antibody titer $\geq 4$, n (%) | 246(96.85) | 241(96.79) | 0.9678 |
| 95% CI (%) | 93.89-98.63 | 93.77-98.60 |  |
| Pre-booster antibody GMT<sup>[1]</sup> (95%CI) | 39.19(34.27-44.82) | 40.35(35.05-46.45) | 0.7689 |
| Post-booster antibody GMT (95%CI) | 983.36(852.25-1134.64) | 117.33(103.07-133.55) | <0.0001 |
| Ratio of GMT between two groups(95%CI)<sup>[2]</sup> | 8.38(6.91-10.16) |  |  |
| Post-booster adjusted antibody GMT (95%CI)<sup>[3]</sup> | 939.60(742.60-1188.86) | 110.57(87.09-140.38) | <0.0001 |
| Ratio of adjusted GMT between two groups(95%CI)<sup>[3]</sup> | 8.50(7.15-10.10) |  |  |
| Rate of 4-fold rise<sup>[4]</sup>, n (%) | 240(94.49) | 104(41.77) | <0.0001 |
| 95% CI (%) | 90.92-96.95 | 35.57-48.16 |  |
| Rate difference between two groups (% , 95%CI<sup>[5]</sup>) | 52.76(46.02-59.51) |  |  |
| Post-booster antibody GMT fold rise (95%CI) | 25.09(21.17-29.73) | 2.91(2.60-3.25) | <0.0001 |
| **All subjects (Excluding participants with positive or weak positive COVID-19 PCR test)** |  |  |  |
| N(missing) | 231(1)<sup>[6]</sup> | 226(0) |  |
| Pre-booster antibody titer $\geq 4$, n (%) | 224(97.39) | 218(96.46) | 0.5644 |
| 95% CI (%) | 94.41-99.04 | 93.14-98.46 |  |
| Pre-booster antibody GMT<sup>[1]</sup> (95%CI) | 40.35(35.15-46.33) | 40.53(34.98-46.97) | 0.9649 |
| Post-booster antibody GMT (95%CI) | 985.80(847.56-1146.58) | 118.60(103.61-135.75) | <0.0001 |
| Ratio of GMT between two groups(95%CI)<sup>[2]</sup> | 8.31(6.79-10.18) |  |  |
| Post-booster adjusted antibody GMT (95%CI)<sup>[3]</sup> | 914.93(713.58-1173.11) | 109.55(84.81-141.50) | <0.0001 |
| Ratio of adjusted GMT between two groups(95%CI)<sup>[3]</sup> | 8.35(6.95-10.03) |  |  |
|  | NVSI-06-09 | BBIBP-CorV | *p* value |
| Rate of 4-fold rise<sup>[4]</sup>, n (%) | 217(94.35) | 98(43.36) | <0.0001 |
| 95% CI (%) | 90.53-96.96 | 36.81-50.10 |  |
| Rate difference between two groups (% , 95% CI<sup>[5]</sup>) | 51.03(43.89-58.16) |  |  |
| Post-booster antibody GMT fold rise (95% CI ) | 24.43(20.44-29.20) | 2.93(2.59-3.30) | <0.0001 |
| **Subjects who have received 2 doses of inactivated vaccine previously** |  |  |  |
| N(missing) | 11(0) | 9(0) |  |
| Pre-booster antibody titer ≥4, n (%) | 11(100.00) | 9(100.00) | 1.0000 |
| 95% CI (%) | 71.51-100.00 | 66.37-100.00 |  |
| Pre-booster antibody GMT<sup>[1]</sup> (95% CI) | 28.69(16.55-49.72) | 80.17(53.95-119.13) | 0.0043 |
| Post-booster antibody GMT (95% CI) | 724.52(416.59-1260.06) | 153.28(79.97-293.80) | 0.0006 |
| Ratio of GMT between two groups(95% CI)<sup>[2]</sup> | 4.73(2.15-10.39) |  |  |
| Post-booster adjusted antibody GMT (95% CI)<sup>[3]</sup> | 747.89(405.03-1380.97) | 147.44(73.68-295.04) | 0.0039 |
| Ratio of adjusted GMT between two groups(95% CI)<sup>[3]</sup> | 5.07(1.82-14.14) |  |  |
| Rate of 4-fold rise<sup>[4]</sup>, n (%) | 11(100.0) | 2(22.22) | 0.0004 |
| 95% CI (%) | 71.51-100.00 | 2.81-60.01 |  |
| Rate difference between two groups (% , 95% CI<sup>[5]</sup>) | 77.78(50.62-100.00) |  |  |
| Post-booster antibody GMT fold rise (95% CI ) | 25.26(11.26-56.65) | 1.91(1.00-3.65) | <0.0001 |
| **Subjects who have received 3 doses of inactivated vaccine previously** |  |  |  |
| N(missing) | 244(1)<sup>[6]</sup> | 240(0) |  |
| Pre-booster antibody titer ≥4, n (%) | 235(96.71) | 232(96.67) | 0.9798 |
| 95% CI (%) | 93.62-98.57 | 93.54-98.55 |  |
| Pre-booster antibody GMT<sup>[1]</sup> (95% CI) | 39.75(34.61-45.66) | 39.32(34.03-45.44) | 0.9149 |
| Post-booster antibody GMT (95% CI) | 997.05(859.99-1155.95) | 116.16(101.71-132.66) | <0.0001 |
| Ratio of GMT between two groups(95% CI)<sup>[2]</sup> | 8.58(7.04-10.47) |  |  |
|  | NVSI-06-09 | BBIBP-CorV | *p* value |
| Post-booster adjusted antibody GMT (95%CI)<sup>[3]</sup> | 994.62(878.04-1126.68) | 116.44(102.72-132.01) | <0.0001 |
| Ratio of adjusted GMT between two groups(95%CI)<sup>[3]</sup> | 8.54(7.16-10.19) |  |  |
| Rate of 4-fold rise<sup>[4]</sup>, n (%) | 229(94.24) | 102(42.50) | <0.0001 |
| 95%CI (%) | 90.52-96.81 | 36.16-49.02 |  |
| Rate difference between two groups (% , 95%CI<sup>[5]</sup>) | 51.74(44.83-58.65) |  |  |
| Post-booster antibody GMT fold rise (95%CI) | 25.08(21.06-29.88) | 2.95(2.64-3.30) | <0.0001 |

Notes: [1] GMT represent geometric mean titer.
[2] The ratio of GMT between two groups was calculated by “NVSI-06-09/ BBIBP-CorV”.
[3] Covariance analysis with least square method was used to calculate the adjusted GMT and the corresponding *p* value.
[4] Rate of 4-fold rise was defined as percentage of participants with a $\geq 4$ -fold rise in neutralizing antibody titer from baseline.
[5] Rate difference=(NVSI-06-09)-(BBIBP-CorV). Rate difference and 95%CI were estimated by CMH method considering stratification factors.
[6] The serum sample of one subject was not tested due to contamination.

**Table S5.**
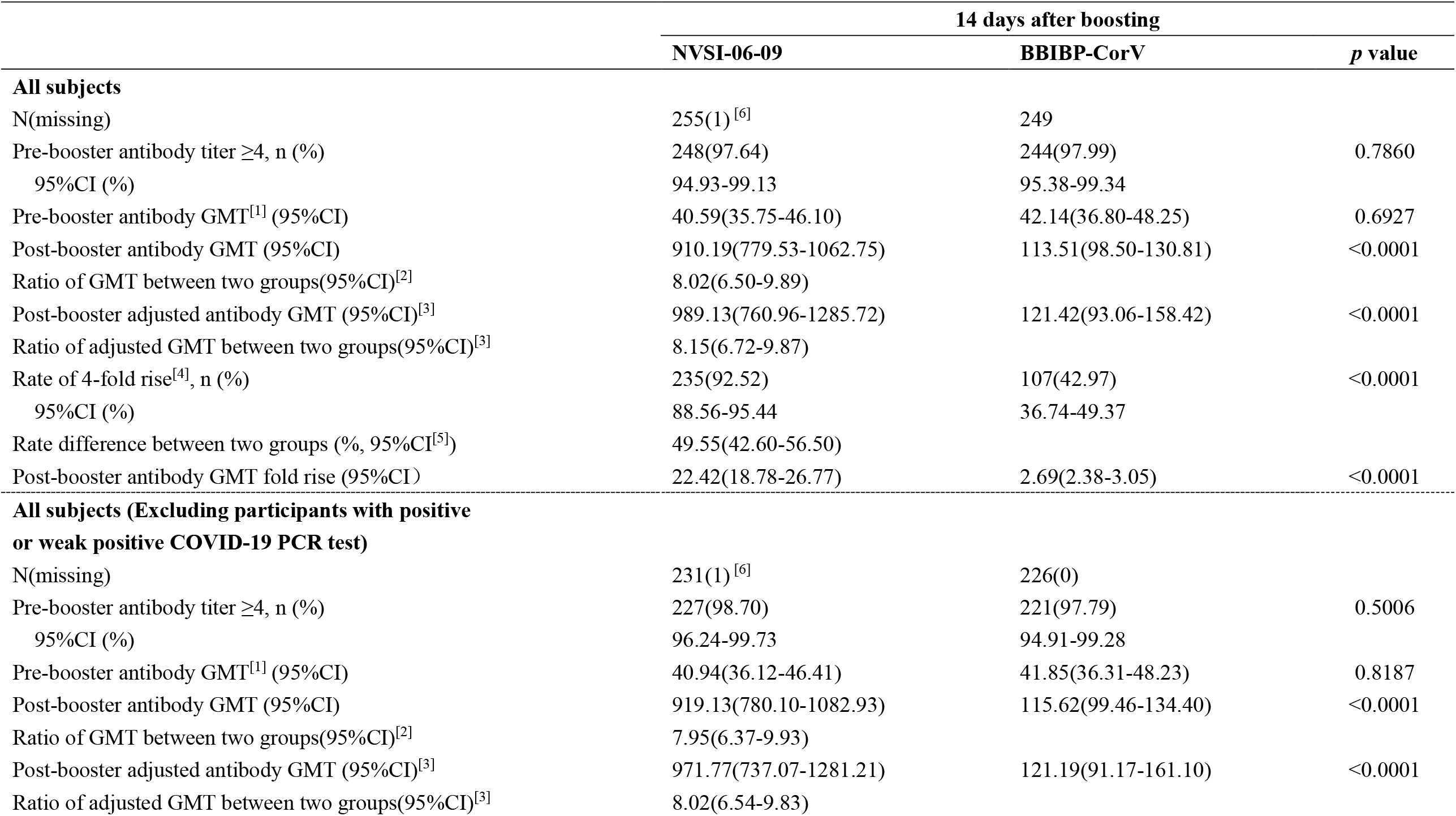
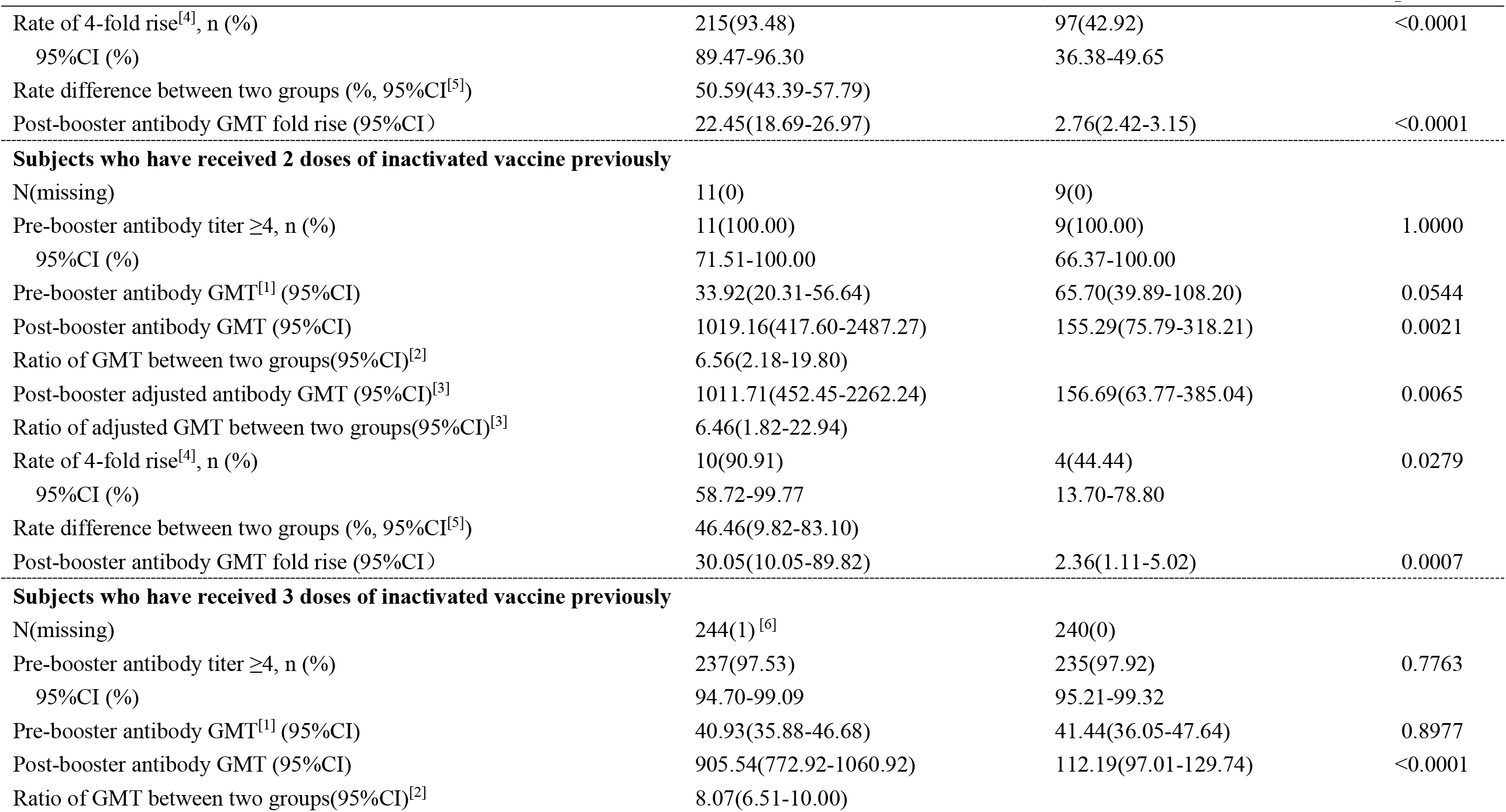
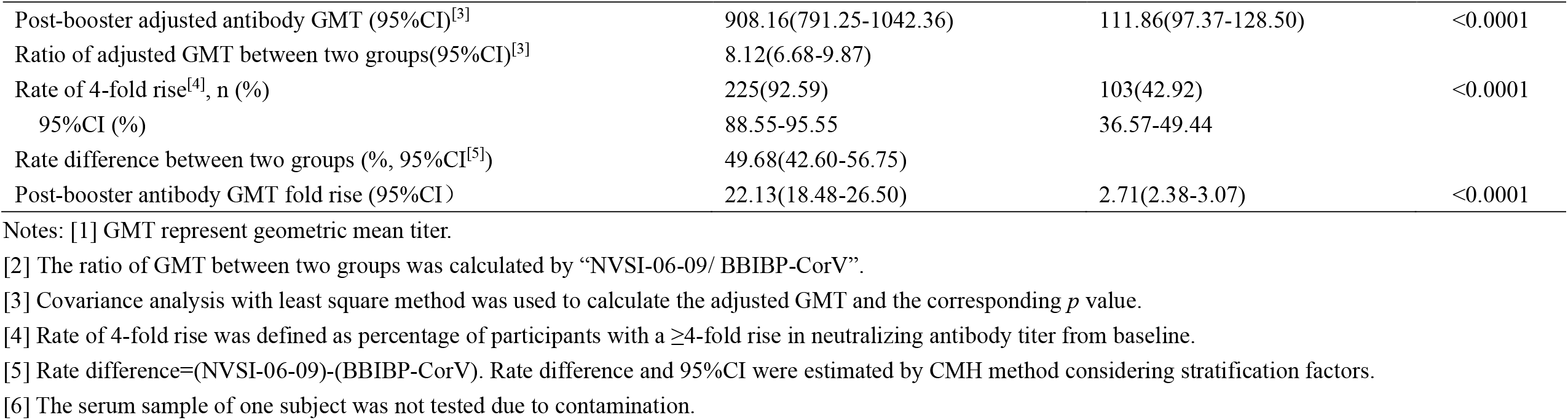
Live-virus neutralizing antibody response against Omicron (BA.4) variant (PPS)

**Table S6.** Live-virus neutralizing antibody response against Omicron (BA.5) variant (PPS)

|  | 14 days after boosting |  |  |
| --- | --- | --- | --- |
|  | NVSI-06-09 | BBIBP-CorV | *p* value |
| **All subjects** |  |  |  |
| N(missing) | 255(1)<sup>[6]</sup> | 249(0) |  |
| Pre-booster antibody titer $\geq 4$, n (%) | 248(97.64) | 238(95.58) | 0.2021 |
| 95% CI (%) | 94.93-99.13 | 92.23-97.77 |  |
| Pre-booster antibody GMT<sup>[1]</sup> (95%CI) | 36.95(32.38-42.16) | 34.98(30.53-40.09) | 0.5707 |
| Post-booster antibody GMT (95%CI) | 999.85(872.41-1145.90) | 165.39(142.99-191.30) | <0.0001 |
| Ratio of GMT between two groups(95%CI)<sup>[2]</sup> | 6.05(4.96-7.38) |  |  |
| Post-booster adjusted antibody GMT (95%CI)<sup>[3]</sup> | 910.53(708.86-1169.57) | 153.82(119.34-198.27) | <0.0001 |
| Ratio of adjusted GMT between two groups(95%CI)<sup>[3]</sup> | 5.92(4.93-7.11) |  |  |
| Rate of 4-fold rise<sup>[4]</sup>, n (%) | 241(94.88) | 147(59.04) | <0.0001 |
| 95% CI (%) | 91.41-97.25 | 52.65-65.20 |  |
| Rate difference between two groups (% , 95%CI<sup>[5]</sup>) | 35.95(29.28-42.62) |  |  |
| Post-booster antibody GMT fold rise (95%CI) | 27.06(23.04-31.78) | 4.73(4.10-5.45) | <0.0001 |
| **All subjects (Excluding participants with positive or weak positive COVID-19 PCR test)** |  |  |  |
| N(missing) | 231(1)<sup>[6]</sup> | 226(0) |  |
| Pre-booster antibody titer $\geq 4$, n (%) | 226(98.26) | 215(95.13) | 0.0611 |
| 95% CI (%) | 95.61-99.52 | 91.46-97.55 |  |
| Pre-booster antibody GMT<sup>[1]</sup> (95%CI) | 37.60(32.85-43.03) | 34.73(30.03-40.15) | 0.4300 |
| Post-booster antibody GMT (95%CI) | 999.29(864.82-1154.67) | 166.48(142.50-194.49) | <0.0001 |
| Ratio of GMT between two groups(95%CI)<sup>[2]</sup> | 6.00(4.86-7.42) |  |  |
| Post-booster adjusted antibody GMT (95%CI)<sup>[3]</sup> | 902.05(692.51-1174.98) | 154.95(118.03-203.42) | <0.0001 |
| Ratio of adjusted GMT between two groups(95%CI)<sup>[3]</sup> | 5.82(4.79-7.07) |  |  |
|  | NVSI-06-09 | BBIBP-CorV | *p* value |
| Rate of 4-fold rise<sup>[4]</sup>, n (%) | 218(94.78) | 136(60.18) | <0.0001 |
| 95% CI (%) | 91.06-97.28 | 53.47-66.61 |  |
| Rate difference between two groups (% , 95% CI<sup>[5]</sup>) | 34.77(27.78-41.76) |  |  |
| Post-booster antibody GMT fold rise (95% CI) | 26.58(22.48-31.42) | 4.79(4.12-5.57) | <0.0001 |
| **Subjects who have received 2 doses of inactivated vaccine previously** |  |  |  |
| N(missing) | 11(0) | 9(0) |  |
| Pre-booster antibody titer ≥4, n (%) | 11(100.00) | 9(100.00) | 1.0000 |
| 95% CI (%) | 71.51-100.00 | 66.37-100.00 |  |
| Pre-booster antibody GMT<sup>[1]</sup> (95% CI) | 28.08(17.30-45.56) | 61.63(30.32-125.26) | 0.0461 |
| Post-booster antibody GMT (95% CI) | 908.15(415.39-1985.47) | 139.04(72.29-267.42) | 0.0008 |
| Ratio of GMT between two groups(95% CI)<sup>[2]</sup> | 6.53(2.45-17.39) |  |  |
| Post-booster adjusted antibody GMT (95% CI)<sup>[3]</sup> | 950.99(466.28-1939.58) | 131.43(59.24-291.59) | 0.0018 |
| Ratio of adjusted GMT between two groups(95% CI)<sup>[3]</sup> | 7.24(2.34-22.34) |  |  |
| Rate of 4-fold rise<sup>[4]</sup>, n (%) | 10(90.91) | 3(33.33) | 0.0089 |
| 95% CI (%) | 58.72-99.77 | 7.49-70.07 |  |
| Rate difference between two groups (% , 95% CI<sup>[5]</sup>) | 57.58(22.40-92.75) |  |  |
| Post-booster antibody GMT fold rise (95% CI) | 32.34(11.97-87.37) | 2.26(1.14-4.47) | 0.0002 |
| **Subjects who have received 3 doses of inactivated vaccine previously** |  |  |  |
| N(missing) | 244(1)<sup>[6]</sup> | 240(0) |  |
| Pre-booster antibody titer ≥4, n (%) | 237(97.53) | 229(95.42) | 0.2074 |
| 95% CI (%) | 94.70-99.09 | 91.95-97.69 |  |
| Pre-booster antibody GMT<sup>[1]</sup> (95% CI) | 37.41(32.64-42.88) | 34.25(29.81-39.35) | 0.3724 |
| Post-booster antibody GMT (95% CI) | 1004.21(873.58-1154.38) | 166.47(143.33-193.34) | <0.0001 |
| Ratio of GMT between two groups(95% CI)<sup>[2]</sup> | 6.03(4.92-7.40) |  |  |
|  | NVSI-06-09 | BBIBP-CorV | *p* value |
| Post-booster adjusted antibody GMT (95%CI)<sup>[3]</sup> | 985.96(864.03-1125.11) | 169.59(148.49-193.68) | <0.0001 |
| Ratio of adjusted GMT between two groups(95%CI)<sup>[3]</sup> | 5.81(4.82-7.01) |  |  |
| Rate of 4-fold rise<sup>[4]</sup>, n (%) | 231(95.06) | 144(60.00) | <0.0001 |
| 95%CI (%) | 91.53-97.42 | 53.50-66.25 |  |
| Rate difference between two groups (% , 95%CI<sup>[5]</sup>) | 35.06(28.29-41.83) |  |  |
| Post-booster antibody GMT fold rise (95%CI) | 26.84(22.79-31.61) | 4.86(4.20-5.62) | <0.0001 |

Notes: [1] GMT represent geometric mean titer.
[2] The ratio of GMT between two groups was calculated by “NVSI-06-09/ BBIBP-CorV”.
[3] Covariance analysis with least square method was used to calculate the adjusted GMT and the corresponding *p* value.
[4] Rate of 4-fold rise was defined as percentage of participants with a $\geq 4$ -fold rise in neutralizing antibody titer from baseline.
[5] Rate difference=(NVSI-06-09)-(BBIBP-CorV). Rate difference and 95%CI were estimated by CMH method considering stratification factors.
[6] The serum sample of one subject was not tested due to contamination.

**Table S7.** Live-virus neutralizing antibody responses against Beta and Delta variants.

|  | **NVSI-06-09** | **BBIBP-CorV** | **GMT ratio and 95%CI** | ***p* value** |
| --- | --- | --- | --- | --- |
| **Beta strain** |  |  |  |  |
| n | 99 | 100 |  |  |
| GMT (95%CI) | 3075.59(2393.00-3952.89) | 465.69(376.75-575.63) | 6.60(4.77-9.15) | <0.0001 |
| **Delta strain** |  |  |  |  |
| n | 99 | 100 |  |  |
| GMT (95%CI) | 2831.50(2200.42-3643.58) | 395.05(316.82-492.58) | 7.17(5.14-10.00) | <0.0001 |

Notes: “n” is the number of serum samples used in the test.

